# Multimodal Prediction of Primary Open-Angle Glaucoma Using Polygenic Risk Scores and Clinical Features in a High-Risk African Ancestry Cohort

**DOI:** 10.1101/2025.09.27.25336797

**Authors:** Yan Zhu, Aude Benigne Ikuzwe Sindikubwabo, Yuki Bradford, Lannawill Caruth, Rebecca Salowe, Kenneth Pham, Laxmi Moksha, Vrathasha Vrathasha, Roy Lee, Mina Halimitabrizi, Isabel Di Rosa, Leila Ghaffari, Jie He, Penn Medicine BioBank, Regeneron Genetics Center, Marylyn D. Ritchie, Joan M. O’Brien, Shefali Setia-Verma

## Abstract

Primary open-angle glaucoma (POAG) disproportionately affects individuals of African ancestry, for whom risk prediction tools remain limited. We evaluated four polygenic risk scores (PGS), two genome-wide and two curated, weighted by African ancestry effect sizes. Used alone, the scores showed modest discrimination. Age and sex were strongly predictive; adding PGS616 reached a cross-validated AUC of 0.713 in the training cohort (N = 271), comparable to the demographic model. In the Penn Medicine BioBank (N = 9,084) the model reached AUC = 0.727, similar to 0.729 for age and sex alone, with comparable case detection at fixed specificity. In a suspect cohort (N = 1,013), predicted risk tracked cup-to-disc ratio and nerve fiber layer thinning, an association explained by age and sex. These results establish a demographic benchmark for evaluating ancestry-matched PGS in POAG.

**HIGHLIGHTS:**

- Age and sex alone predict POAG well in a high-risk African ancestry cohort
- Four ancestry-matched PGS show modest standalone discrimination in this cohort
- Adding PGS616 to age and sex gives comparable AUC in both cohorts (primary MLP)
- Predicted risk tracks optic nerve damage in suspects, driven by age and sex

## INTRODUCTION

Glaucoma is the leading cause of irreversible blindness worldwide, affecting over 70 million individuals and projected to rise dramatically with the aging population.^1,2^ Primary open-angle glaucoma (POAG), the most prevalent form of this disease, accounts for approximately 74% of all diagnosed glaucoma cases.^3^ POAG is characterized by progressive degeneration of the optic nerve, often accompanied by elevated intraocular pressure (IOP), thinning of the retinal nerve fiber layer (RNFL), structural changes in the optic disc such as increased cup-to-disc ratio (CDR), and corresponding loss of visual field.^4–7^ Importantly, individuals of African ancestry are disproportionately affected by POAG, experiencing earlier onset, more rapid progression, and up to five-fold higher risk of blindness compared to those of European ancestry.^8,9^ Despite these known disparities, early detection and risk stratification in this high-risk population remain inadequate, with over 50% of cases remaining undiagnosed.^10,11^ This underscores an urgent need for precision tools that can support earlier and more accurate diagnosis.^12^

In recent years, advances in genomics have enabled the development of polygenic risk scores (PGS), which aggregate the effects of multiple genetic variants to estimate an individual’s inherited susceptibility to complex diseases.^13–15^ These tools have shown promise in a range of common diseases, including type 2 diabetes,^16,17^ coronary heart disease,^18,19^ and breast cancer,^20–22^ and are just beginning to gain traction in ophthalmic diseases with a strong genetic component, such as POAG.^23,24^ However, there are several major limitations to existing PGS for POAG. First, the majority of existing risk scores are trained in predominantly European ancestry cohorts and often lack transferability to other ancestries.^25,26^ Therefore, PGS built from carefully selected, biologically validated, and ancestry-matched variants may offer a more interpretable and potentially more reliable approach for glaucoma risk prediction.^22,27,28^ Additionally, while existing PGS models reflect a person’s inherited susceptibility to the disease, these risk scores do not consistently integrate current disease status or severity. In contrast, routinely measured clinical features, such as IOP, RNFL thickness, and CDR, are valuable diagnostic indicators that capture disease after structural or functional damage has already occurred.^29^ However, key clinical features such as IOP are pharmacologically modifiable and may therefore not fully reflect underlying disease biology, particularly in patients receiving IOP-lowering therapy. To solve this, integrating these complementary data types could yield more accurate and equitable risk prediction models for POAG.^25^

Artificial intelligence (AI), particularly supervised machine learning (ML), offers a powerful framework to harness the complexity of multidimensional health data.^30,31^ In contrast to traditional regression-based risk models, ML algorithms can capture non-linear interactions among diverse inputs and automatically learn relevant feature representations.^32,33^ In ophthalmology, ML techniques have been successfully employed to analyze fundus photographs, segment optic nerve head structures, and quantify RNFL thickness using OCT scans.^34–36^ These models can detect structural changes that are often imperceptible to the human eye, providing a critical advantage in screening and disease monitoring.^37–39^ However, relatively few studies have examined the utility of integrating genetic risk, particularly PGS, with ocular phenotypes in large, ancestrally diverse POAG cohorts.^40,41^ Moreover, no prior work has systematically compared multiple ML models across different combinations of phenotypic and genetic features in the most overaffected African ancestry cohort. This gap highlights the need to evaluate whether ancestry-matched genetic scores, especially loci-focused PGS, can provide added predictive value beyond standard phenotypes in the context of ML-based models.

In this study, we evaluated ML models incorporating PGS for POAG prediction in the Primary Open-Angle African Ancestry Glaucoma Genetics (POAAGG) cohort, which is a large and deeply phenotyped African ancestry dataset.^9,42,43^ We constructed four complementary PGS, including two genome-wide scores (POAAGG PGS and MEGA PGS) that capture broad polygenic effects,^44^ and two loci-based scores (PGS526 and PGS616) derived from curated multi-ancestry glaucoma loci and weighted using African ancestry effect sizes.^14,45–49^ Using these PGS, we trained four supervised learning models in a POAAGG training cohort of 271 subjects, including logistic regression (LR),^50,51^ multilayer perceptron (MLP),^52^ random forest (RF),^53–55^ and support vector machine (SVM).^56–58^ These models were evaluated for performance and generalizability in an external validation cohort of African ancestry participants in the Penn Medicine BioBank (PMBB) (N=9,084) and were tested in an independent suspect cohort of 1,013 POAAGG subjects.^59^ Each PGS was integrated into ML models alongside demographic features, and in a separate analysis alongside inter-eye clinical asymmetry, to assess whether genetic information could improve predictive accuracy across the models.

## RESULTS

### Study Design and Data Integration Framework

We developed a structured pipeline that integrates PGS generation, model training, and biological validation (Figure 1). Four PGS were constructed using two complementary strategies: genome-wide PGS derived from African ancestry GWAS via PRS-CS^44^ (POAAGG PGS and MEGA PGS) and loci-based PGS built from six multi-ancestry glaucoma GWAS^14,45–49^ weighted by African ancestry-specific effect sizes (PGS526 and PGS616).

**Figure 1.**
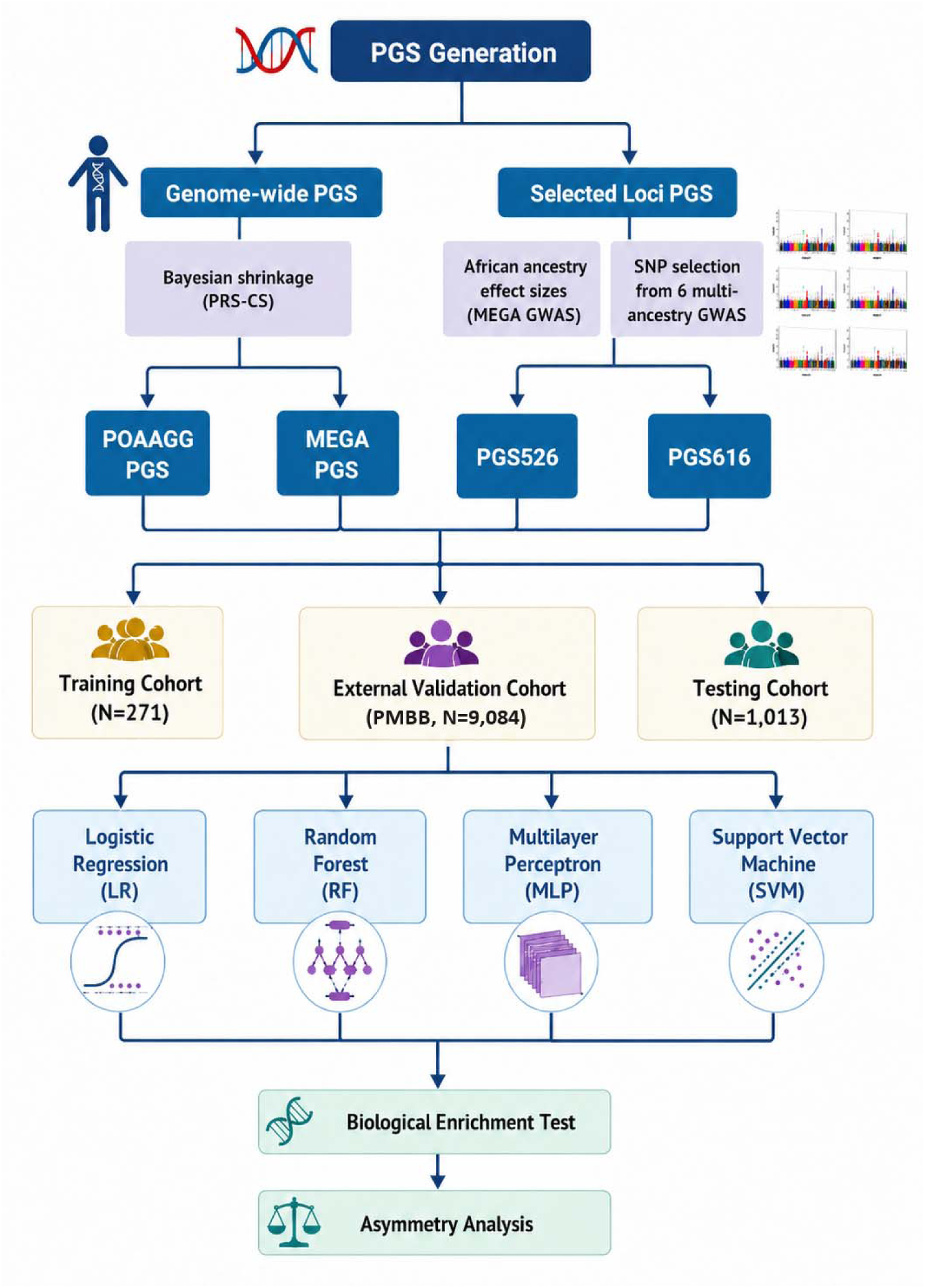
Study Design and Data Integration Framework. Overview of the analytical pipeline integrating polygenic risk score (PGS) generation, machine learning model training, and biological validation. Four PGS were developed using two complementary strategies: genome-wide Bayesian shrinkage via PRS-CS yielding the POAAGG PGS and MEGA PGS, and curated locus-based selection of SNPs from six multi-ancestry GWAS weighted by African ancestry effect sizes (MEGA GWAS for PGS616; POAAGG MTAG for PGS526), yielding PGS526 (526 SNPs) and PGS616 (616 SNPs). All four PGS were evaluated across three cohorts: a POAAGG case-control training cohort (N = 271; 128 cases, 143 controls; mean age = 65; 160 females, 111 males), an independent external validation cohort from the Penn Medicine BioBank (PMBB; N = 9,084 African ancestry individuals), and an independent POAAGG suspect cohort (N = 1,013; mean age = 63; 671 females, 342 males). Four supervised classifiers were applied: logistic regression (LR), random forest (RF), multilayer perceptron (MLP), and support vector machine (SVM). Trained models were applied without retraining to the suspect cohort for biological enrichment testing and inter-eye asymmetry analysis.

Model training was conducted in a training cohort of 271 subjects (128 cases and 143 controls; mean age = 65 years; 160 females, 111 males, summarized in Table S1). The training set showed clear phenotypic separation between cases and controls. Cases exhibited significantly higher baseline IOP (21.70 ± 6.71 mmHg vs. 14.73 ± 2.96 mmHg; Welch’s t-test p = 5.44 × 10⁻ ²¹), larger baseline CDR (0.76 ± 0.14 vs. 0.32 ± 0.11; p = 4.86 × 10⁻ □□), and thinner baseline RNFL (69.62 ± 9.48 µm vs. 96.84 ± 8.53 µm; p = 4.01 × 10⁻ □□).

To evaluate model generalizability, predictions were validated in an external cohort from the PMBB, a large health system-based biorepository at the University of Pennsylvania.^59^ We included PMBB participants with genetically inferred African ancestry aged ≥35 years, with POAG cases defined by harmonized International Classification of Diseases (ICD)-based criteria (ICD-9: 365.10/365.11; ICD-10: H40.11 variants) and controls defined as individuals with no evidence of glaucoma diagnosis. This yielded an external validation cohort of 9,084 participants (See *STAR Methods* for details).

The trained models were also tested in an independent suspect cohort of 1,013 subjects (mean age = 63 years; 671 females, 342 males).^60^ The suspect group represents individuals with early or subclinical glaucoma features, exhibiting intermediate clinical profiles for IOP (20.16 ± 6.02 mmHg), CDR (0.59 ± 0.15), and RNFL thickness (86.35 ± 12.48 µm). Because definitive case and control labels are unavailable in this suspect population, model performance was assessed using biological enrichment analyses, testing whether higher model-predicted risk aligned with established ocular phenotypes (IOP, CDR, RNFL thickness). In addition, given that early POAG frequently presents with inter-eye asymmetry, we further evaluated whether PGS-related models could capture clinically meaningful asymmetry patterns, as an additional, though not fully independent, check on the relation between ancestry-informed genetic risk and clinically important glaucoma phenotypes.

### Standalone Polygenic Scores Show Modest Discrimination of POAG

To capture the inherited component of POAG risk, we constructed four PGS using two complementary strategies (See *STAR Methods*). The first strategy employed a genome-wide approach aggregating risk across millions of common variants, yielding two scores: the POAAGG PGS (based on the POAAGG cohort GWAS) and the MEGA PGS (Multi-Ethnic Glaucoma Analysis; based on a larger African ancestry mega-analysis).^14,44^ The second strategy used biologically curated, loci-based scores derived from genome-wide significant SNPs across six large glaucoma GWAS,^14,45–49^ weighted by African ancestry effect sizes: PGS616, weighted by case–control effect sizes from the MEGA GWAS, and PGS526, the subset of those variants weighted by effect sizes from a multi-trait analysis of IOP, CDR and RNFL endophenotypes. A complete list of SNPs and weights is provided in Table S2.

To assess genomic coverage, we examined the chromosomal distribution of variants included in the two loci-based PGS, highlighting their overlap and complementarity (Figure 2A; Figure S1 for full circos layout). The bar chart illustrates the number of genetic variants per chromosome included in PGS526 and PGS616, demonstrating their broad genomic coverage across all autosomes. PGS526 is a strict subset of PGS616: all 526 variants are contained in the 616-variant panel, and the 90 additional variants are disease-status loci without a usable MTAG weight. The two scores therefore differ principally in the source of their effect-size weights rather than in variant membership (Table S3). We next evaluated how these variant-level differences translated to score-level behavior across cohorts. Figure 2B shows the standardized PGS distributions in training cases, training controls and suspects; for every score the distributions overlap substantially, consistent with the near-chance standalone discrimination described below.

**Figure 2.**
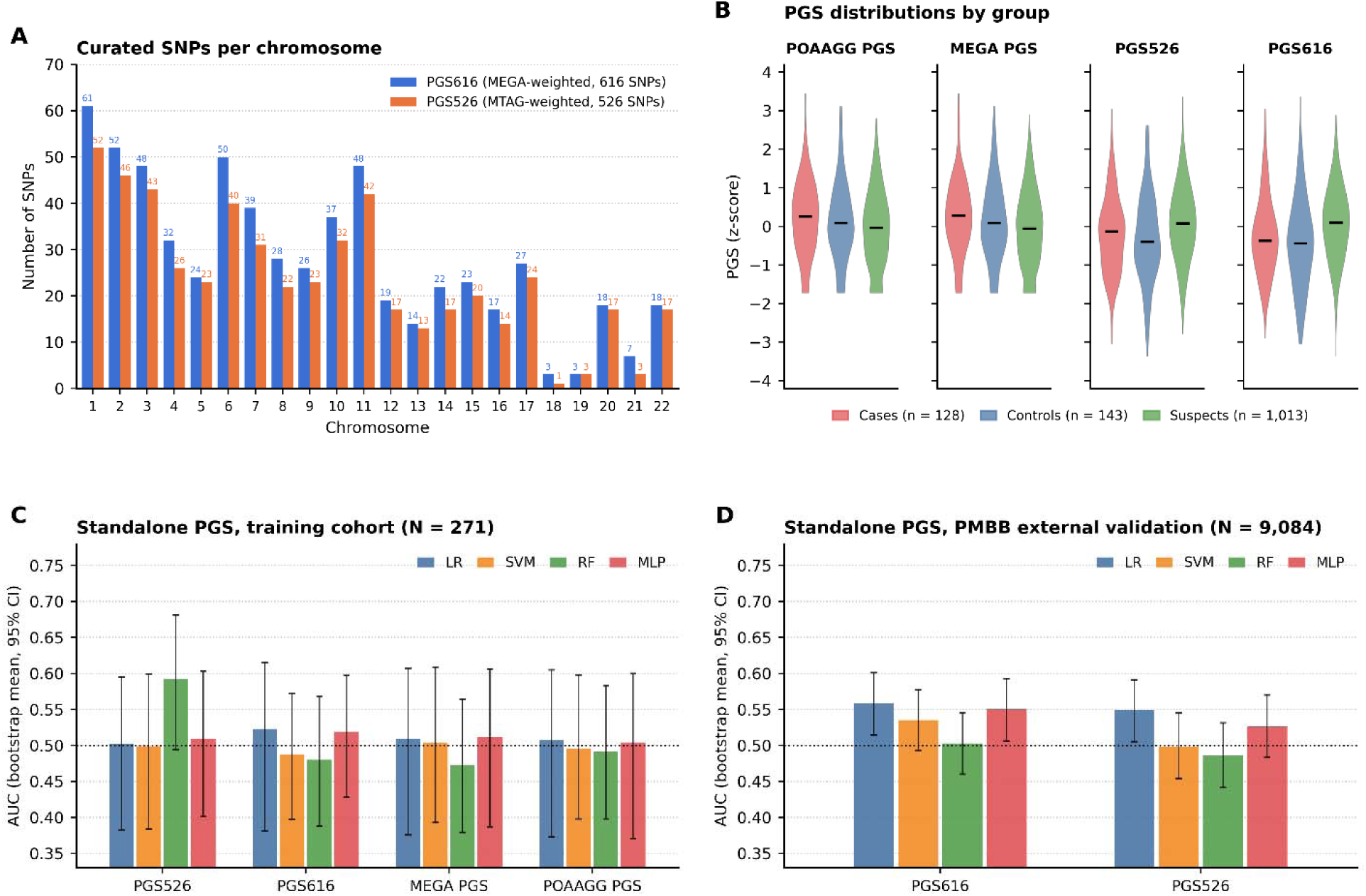
Standalone PGS Performance in Training and External Validation Cohorts. (A) Number of SNPs on each chromosome in the two curated loci-based PGS panels, PGS616 (MEGA-weighted, 616 SNPs; blue) and PGS526 (MTAG-weighted, 526 SNPs; orange), shown as side-by-side bars with the count above each bar. (B) Violin plots of the four PGS in training cases (n = 128; red), training controls (n = 143; blue) and suspects (n = 1,013; green). Each score was rank-based inverse normal transformed across the 1,284 POAAGG participants and z-scored for display; horizontal bars mark group medians. (C) Training cohort (N = 271) area under the receiver operating characteristic curve (AUC) for four classifiers, logistic regression (LR), support vector machine (SVM), random forest (RF) and multilayer perceptron (MLP), each trained on a single PGS. Bars show the participant-level bootstrap mean AUC and error bars the 95% percentile confidence intervals (B = 2,000 replicates); the dotted line marks AUC = 0.5. All intervals include 0.5; cross-validation means are given in Table S5. (D) External validation AUC in the Penn Medicine BioBank (PMBB; N = 9,084 with both PGS616 and PGS526 available) for the four classifiers trained on PGS616 or PGS526 in the POAAGG training cohort and applied without refitting. Bars show the bootstrap mean AUC and error bars the 95% confidence intervals (B = 2,000); the dotted line marks AUC = 0.5. AUCs ranged from 0.486 to 0.559. See also Tables S2, S4, S5 and S15.

When used as standalone features, all four PGS demonstrated modest predictive performance in the training cohort (Figure 2C), with modest average AUC values, motivating their evaluation within combined models. Average AUC values across the four machine learning classifiers (LR, RF, MLP, SVM) using 5-fold × 20-repeat stratified cross-validation showed the following ordering, which is not statistically distinguishable (Table S4). PGS526 achieved the highest mean AUC (0.526 ± 0.068) averaged across classifiers, followed by MEGA PGS (0.500 ± 0.072), PGS616 (0.500 ± 0.062), and POAAGG PGS (0.498 ± 0.070). The best individual configuration, RF with PGS526, achieved AUC = 0.598 ± 0.065. Under participant-level bootstrap resampling, the 95% confidence intervals of all four scores included 0.5 (PGS526 0.526, 95% CI 0.451-0.588; PGS616 0.503, 0.431-0.559; MEGA PGS 0.499, 0.419-0.567; POAAGG PGS 0.500, 0.421-0.565), and no curated score was distinguishable from a genome-wide score in a paired contrast (largest difference PGS526 versus MEGA PGS, +0.026, 95% CI-0.077 to +0.127, p = 0.61; Table S4). Curated and genome-wide scores therefore performed comparably. Detailed performance metrics are listed (Table S5), including AUC, accuracy, F1 score, sensitivity, and specificity for all four classifiers. These AUC values are suggestive rather than definitive, and are best interpreted in the context of the multimodal models presented below. To further evaluate polygenic signals in an external cohort, we assessed standalone PGS performance in 9,084 African ancestry participants from PMBB (Figure 2D). PGS616 and PGS526 achieved average AUC values of 0.537 and 0.515, respectively, consistent with training cohort performance.

### Training Cohort Analysis Reveals Small, Classifier-Dependent Changes from Polygenic Risk Integration

In the training cohort (N = 271), we evaluated four supervised classifiers (LR, RF, SVM, and MLP) across twelve feature sets using 5-fold × 20-repeat stratified cross-validation (5×20 CV; 100 model fits per configuration): age only, sex only, a demographic baseline (Base: age + sex), three PC-augmented baselines (Base+PC2, Base+PC5, Base+PC10), four PGS-extended models (Base + each of the four PGS), and two combined PC+PGS models (Base+PC5+PGS526 and Base+PC5+PGS616).

Integrating loci-based PGS into the demographic baseline produced small, classifier-dependent changes in performance; within each classifier, the incremental AUC of Base+PGS616 over the age+sex baseline was negligible for LR (−0.003), SVM (+0.004), and MLP (+0.001), and larger only for RF (+0.068), where the age+sex baseline itself was weakest (0.616) (Table S6, Table S7, Figure 3A–3B, Table S5). The baseline model (age + sex) achieved a cross-classifier mean cross-validated AUC of 0.683, with classifier-specific values ranging from 0.616 (RF) to 0.713 (LR); under participant-level bootstrap resampling the corresponding estimate is 0.680 (95% confidence interval (CI): 0.605–0.748). Throughout, cross-validation means are reported as descriptive summaries and all confidence intervals and p-values derive from the participant-level bootstrap (Table S8). PC-augmented baselines showed a non-monotonic pattern: Base+PC2 gave the highest mean AUC of 0.704 (bootstrap 0.692, 95% CI: 0.616–0.761), with lower values as further components were added (Base+PC5: 0.688; Base+PC10: 0.672; Base+PC20: 0.657, shown in Figure S2 and Table S5), a pattern consistent with overfitting in this modest-sized cohort (N = 271) although the differences are not statistically distinguishable (Base+PC2 versus baseline +0.012, p = 0.61; Base+PC20 versus baseline −0.038, p = 0.36). Among PGS-augmented models, Base+PGS616 gave the highest mean AUC (0.700; bootstrap 0.691, 95% CI: 0.613–0.764; +0.017 over baseline by cross-validation and +0.011, 95% CI −0.038 to +0.048, p = 0.53, by paired bootstrap), followed by Base+PGS526 (0.696; bootstrap 0.684, 95% CI: 0.609–0.757; +0.013), with LR and MLP classifiers showing the largest nominal values (Base+PGS616: LR = 0.710, MLP = 0.713). Feature sets using curated scores reached nominally higher mean AUC than those using genome-wide scores (Base+MEGA PGS: 0.681; Base+POAAGG PGS: 0.685). Combining PCs and PGS (Base+PC5+PGS616: 0.688; Base+PC5+PGS526: 0.682) performed similarly to Base and PGS-only models, suggesting these features capture overlapping variance. Full performance metrics for all twelve feature sets are reported in Table S5. Although Base+PC2 achieved a mean AUC (0.704; bootstrap 0.692, 95% CI: 0.616–0.761) comparable to Base+PGS616 (0.700; bootstrap 0.691, 95% CI: 0.613–0.764), the confidence intervals overlap almost completely, and no feature set differed significantly from the age and sex baseline in the training cohort (Table S8). The lower values obtained with additional components (Base+PC20: 0.657, Figure S2 and Table S5) are consistent with overfitting in this modest-sized cohort, but we do not interpret the ordering of feature sets as evidence of a real difference.

**Figure 3.**
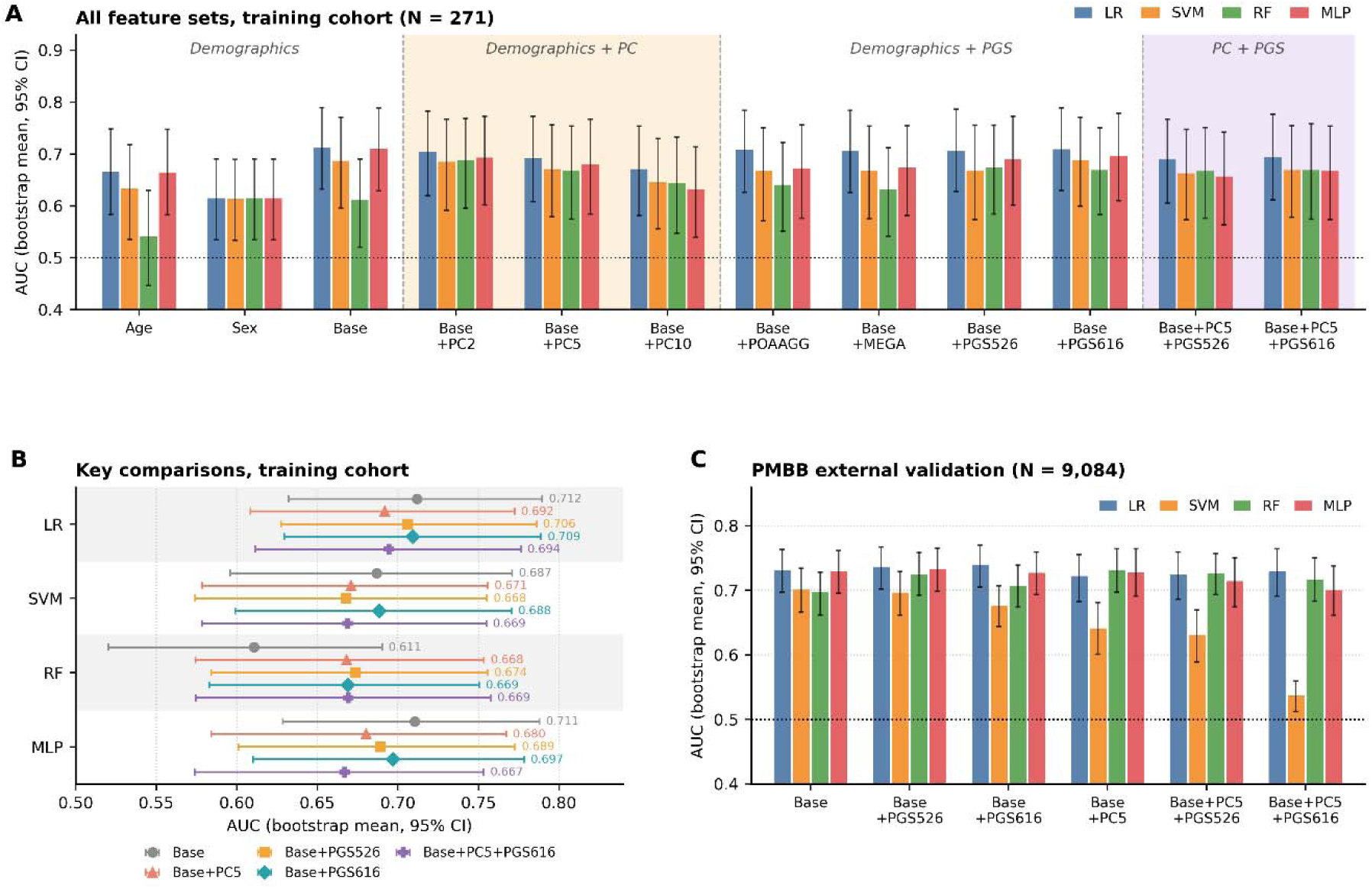
Training and External Validation Performance Across Feature Sets and Classifiers. (A) Area under the receiver operating characteristic curve (AUC; participant-level bootstrap mean and 95% percentil confidence interval [CI], B = 2,000 replicates) for all twelve feature sets across four classifiers, logistic regressio (LR), support vector machine (SVM), random forest (RF) and multilayer perceptron (MLP), in the POAAGG training cohort (N = 271). Feature sets are grouped by category: demographic-only (Age only, Sex only, Base = age + sex), demographic plus ancestry principal components (PCs; Base+PC2 through Base+PC10), polygenic score (PGS)-augmented (Base+POAAGG PGS through Base+PGS616), and combined PC + PGS. The dotted line marks AUC = 0.5. For the primary MLP model, Base (0.711) and Base+PGS616 (0.697) performed comparably, and differences between feature sets are small relative to their confidence intervals. (B) Key configurations (Base, Base+PC5, Base+PGS526, Base+PGS616 and Base+PC5+PGS616) compared within each classifier; points are bootstra means with 95% CIs, and values are printed beside each interval. Base+PGS616 reached a nominally higher AUC than Base+PC5 in every classifier (LR 0.709 vs. 0.692; SVM 0.688 vs. 0.671; RF 0.669 vs. 0.668; MLP 0.697 vs. 0.680), with substantially overlapping intervals. (C) External validation AUC (bootstrap mean and 95% CI, B = 2,000) in the Penn Medicine BioBank (PMBB; N = 9,084 African ancestry participants; 168 cases, 8,916 controls) for six feature sets across the four classifiers; models were trained in the POAAGG training cohort and applied without refitting. The dotted line marks AUC = 0.5. For MLP, Base+PGS616 and the age and sex baseline performed similarly (0.727 versus 0.729). PC-augmented models were lower for some classifiers, most markedly SVM (Base+PC5 0.641 and Base+PC5+PGS616 0.537, versus 0.701 for Base), consistent with ancestry PCs that are not on a common basis across the two cohorts. See also Tables S5, S8 and S15.

To further characterize the relationship between ancestry structure and polygenic risk, we computed Pearson correlations between PC1–PC5 and all four PGS in the training cohort (Figure S2A; full matrix in Table S9). PC3 showed the strongest associations with the curated PGS: PGS616 (r = 0.36, p < 0.001) and PGS526 (r = 0.29, p < 0.001), while correlations with genome-wide PGS were weaker and largely non-significant (all |r| ≤ 0.162). The modest correlations (|r|max = 0.36) indicate that PCs and PGS are only partially overlapping; because a correlation of 0.36 is non-negligible, we do not interpret this as evidence of fully distinct biological signals. Residualizing each PGS on PC1–PC5 removed only a minority of PGS variance (PGS616 retained 85.2% in training and 98.2% in PMBB), indicating the PGS is not merely an ancestry-PC proxy (Table S10). To assess the practical performance impact of PC versus PGS augmentation, we compared participant-level bootstrap AUC across six feature configurations (Figure S2B).

To address model interpretability, we applied SHapley Additive exPlanations (SHAP)^61^ analysis to the MLP classifier across three representative feature configurations: Base, Base+PGS616, and Base+PC5+PGS616 (Figure S3A–B, Table S11). For the primary Base+PGS616 model, age was the most influential feature (mean |SHAP| = 0.127), followed by sex (0.109), while PGS616 contributed less (mean |SHAP| = 0.043). In the SHAP beeswarm plot, older age and male sex generally increased predicted POAG risk, while higher PGS616 values showed a weaker positive contribution. When ancestry principal components (PCs) were included in the Base+PC5+PGS616 model, PC1 became the dominant feature (mean |SHAP| = 0.217), and the relative contribution of PGS616 remained modest. We next evaluated calibration using Brier scores and reliability curves.^62^ In 5-fold calibration analysis of the MLP models, Base+PGS616 showed the lowest Brier score (0.2142), followed by Base (0.2180), whereas Base+PC5+PGS616 showed poorer calibration (0.2304; Figure S3C). In the 5×20 repeated cross-validation analysis across classifiers, MLP calibration showed the same pattern, with Base+PGS616 slightly lower than Base (Brier score 0.2175 vs. 0.2190), a difference small relative to the fold-to-fold standard deviation of about 0.02 (Figure S3D; Table S12).

To assess whether model performance was limited by training sample size, we generated learning curves for three representative feature configurations (Base, Base+PGS616, and Base+PC5+PGS616) using 5-fold × 4-repeat stratified cross-validation, with results averaged across the four classifiers (Figure S4, Table S13). Cross-validation AUC increased with training-set size and did not fully plateau by the maximum available sample size. Base+PGS616 showed the highest average CV AUC at every training size and reached approximately 0.70 at the largest training size, compared with approximately 0.68 for Base and 0.69 for Base+PC5+PGS616. The Base+PC5+PGS616 configuration performed poorly at smaller training sizes and showed a larger train-to-CV gap at full sample size. We further compared training and cross-validation AUCs to assess overfitting (Figure S4B). Base and Base+PGS616 showed moderate train-to-CV gaps (0.065 and 0.083 at full training size), whereas Base+PC5+PGS616 showed the largest gap, particularly at smaller training sizes and persisting at full sample size (0.157).

To evaluate sex-specific heterogeneity, we computed sex-stratified cross-validation AUC within the 5×20 CV framework for male (N = 111, 68 cases) and female (N = 160, 60 cases) subjects separately (Figure S5, Table S14). Female subjects showed higher AUC than males in the Base model (MLP: female = 0.694, male = 0.652), while adding PGS616 narrowed this gap (Base+PGS616 MLP: female = 0.675, male = 0.668). The Base+PC5+PGS616 model reversed this pattern, with a nominally higher AUC in males than females (MLP: female = 0.647, male = 0.726). None of these sex differences was statistically distinguishable under participant-level bootstrap resampling (MLP male minus female: Base −0.045, p = 0.59; Base+PGS616 +0.005, p = 0.96; Base+PC5+PGS616 +0.068, p = 0.47; Figure S5).

In order to evaluate whether these results generalize to an independent population, we applied each trained model to the PMBB external validation cohort (N = 9,084 African ancestry participants; 168 cases, 8,916 controls; Figure 3C; Table S15). The baseline demographic model generalized robustly (mean AUC = 0.714). Adding PGS616 gave a similar value (0.712), and Base+PGS526 was marginally higher (0.723). PC-augmented models showed reduced generalization relative to the base model (Base+PC5: 0.706), and combined PC+PGS models were lower (Base+PC5+PGS526: 0.699; Base+PC5+PGS616: 0.671), suggesting that PC axes from a small training cohort do not transfer robustly to an independent biobank. The strongest external performance was obtained by LR with Base+PGS616 (AUC = 0.739, 95% CI: 0.702–0.771). For the prespecified primary classifier, MLP, age and sex alone reached AUC = 0.729 and the model with PGS616 was comparable (AUC = 0.727; ΔAUC = −0.001, DeLong p = 0.55; Table S6, Table S7). The direction of the increment was classifier-dependent and small throughout: positive for logistic regression (+0.008, p = 0.013), small and non-significant for the random forest (+0.011, p = 0.27) and the multilayer perceptron (−0.001, p = 0.55), and negative for the support vector machine (−0.025, p = 0.008) (Figure S6). To express the increment in screening terms, we carried out an exploratory operating-point analysis of the primary MLP model, setting each model’s decision threshold at 90% and 95% specificity within the external cohort. At 90% specificity the age and sex model detected 26.8% of cases and the PGS-augmented model 26.2% (difference −0.6 percentage points, 95% CI −3.6 to +3.0, p = 0.99); at 95% specificity the values were 10.7% and 12.5% (difference +1.8 percentage points, 95% CI −1.2 to +4.2, p = 0.28). Standardized to the prevalence observed in this cohort (1.85%), these differences correspond to 1.1 fewer and 3.3 more cases detected per 10,000 screened, both within sampling variability. Because the thresholds were set within the validation cohort, they are not independently validated screening thresholds (Table S16).

### Testing Cohort Analysis: Predicted Risk Tracks Structural Damage in an Independent Suspect Cohort, Reflecting Age and Sex

To assess clinical enrichment among individuals without definitive POAG labels, we applied all four trained classifiers across all five feature sets to an independent POAAGG suspect cohort of 1,013 individuals. Full results across all classifier-by-feature-set combinations are shown in Figure S7A–B. For the main figure, we used a single prespecified primary model, MLP with Base+PGS616, to avoid selecting different classifiers for different phenotypes and to maintain consistency with the SHAP and calibration analyses. Each individual in the suspect cohort received a continuous predicted POAG risk score. We tested whether higher predicted risk scores were associated with established clinical indicators of glaucomatous damage, including elevated IOP, enlarged CDR, and thinning of the RNFL. Importantly, these ocular phenotypes were not included as model inputs and were used exclusively for post hoc biological validation. Because definitive case-control labels are unavailable in this suspect population, we assessed Pearson correlations between continuous risk scores and each clinical outcome, and stratified risk into quintiles (Q1–Q5) for visualization of enrichment patterns across increasing predicted POAG risk (Figure 4).

**Figure 4.**
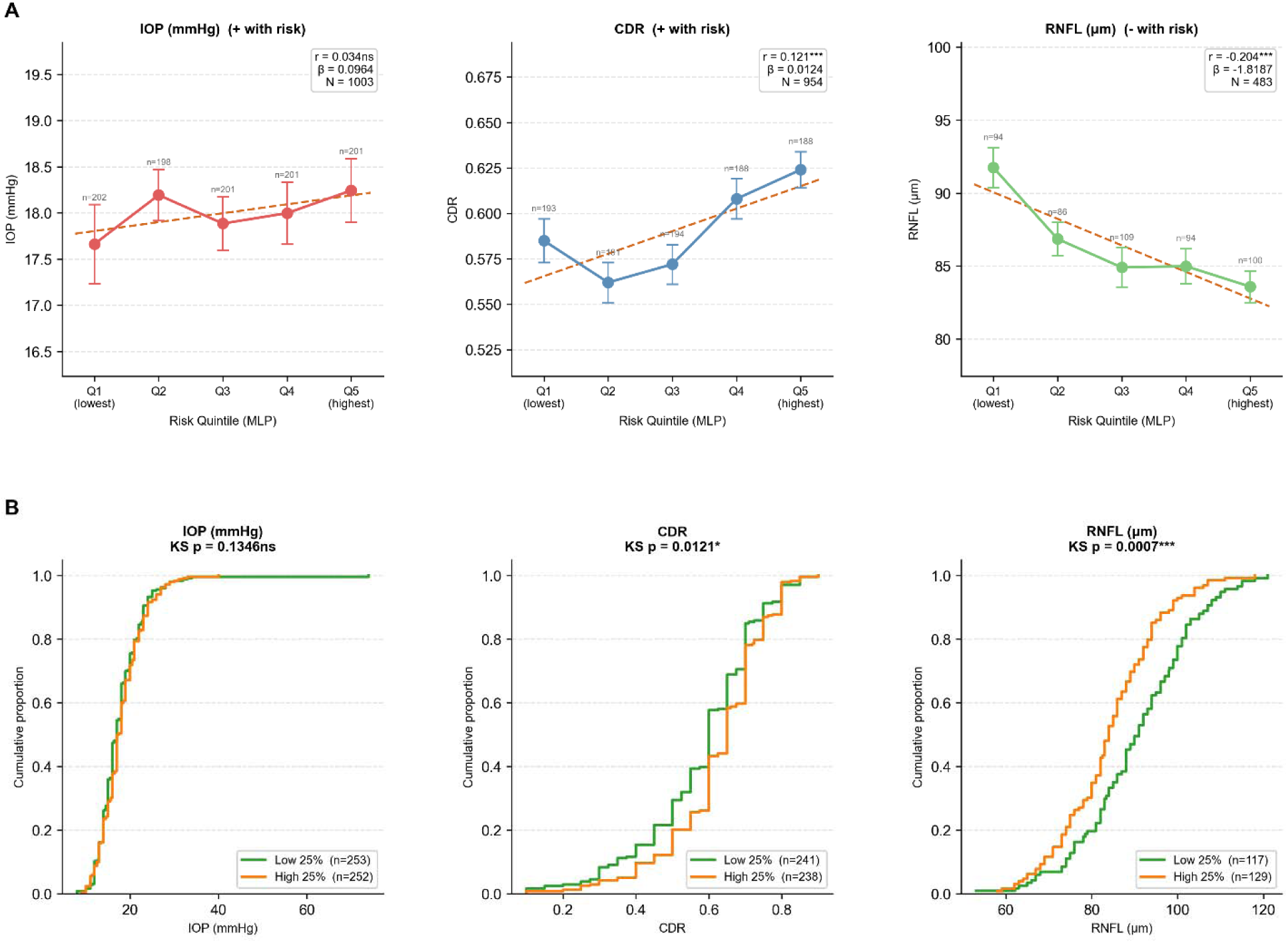
Biological Plausibility and Clinical Enrichment of MLP-Predicted POAG Risk. Biological plausibility validation of multilayer perceptron (MLP)-predicted primary open-angle glaucoma (POAG) risk (Base+PGS616) in the POAAGG suspect cohort (N = 1,013). (A) Quintile stratification plots showing mean ± standard error of the mean (SEM) for intraocular pressure (IOP; mmHg), cup-to-disc ratio (CDR), and retinal nerve fiber layer (RNFL) thickness (µm) across five predicted risk quintiles (Q1 = lowest to Q5 = highest), with Pearson correlation coefficients and p-values. CDR shows a significant positive association (r = 0.121, p < 0.001; N = 954), with a stepwise increase from Q1 (CDR ≈ 0.585) to Q5 (CDR ≈ 0.624). RNFL thickness shows a significant negative association (r = −0.204, p < 0.001; N = 483), declining from Q1 (≈ 91.8 µm) to Q5 (≈ 83.6 µm). IOP does not reach statistical significance (r = 0.034, p = 0.281; N = 1,003), consistent with normal-tension glaucoma biology and potential attenuation by IOP-lowering medications whose use could not be quantified in this dataset. (B) Empirical cumulative distribution functions (ECDFs) comparing phenotype distributions between the bottom 25% (low risk; green) and top 25% (high risk; orange) of predicted POAG risk. Kolmogorov–Smirnov (KS) test p-values are shown. CDR (KS p = 0.0121) and RNFL (KS p = 0.0007) distributions differ significantly between risk groups, indicating that predicted risk tracks structural damage; adjusted analyses (Table S17) show this association reflects age and sex. IOP distributions do not differ significantly (KS p = 0.135).

MLP predicted risk demonstrated significant associations with structural markers of glaucomatous damage (Figure 4A). CDR was significantly positively correlated with predicted risk (r = 0.121, p < 0.001), indicating that suspects with higher predicted POAG risk tended to have larger optic nerve cups, a hallmark of progressive glaucomatous damage. RNFL thickness showed a significant negative correlation (r = −0.204, p < 0.001). Because the predictor contains age and sex, which are themselves related to these phenotypes, we tested the genetic component directly. After adjustment for age and sex, the partial associations of PGS616 with CDR (partial r = 0.025, p = 0.45), RNFL (partial r = 0.032, p = 0.49) and IOP (partial r = 0.029, p = 0.36) were small, and it added less than 0.1% to the variance explained in any phenotype (ΔR² < 0.001; Table S17). Predicted risk from age and sex alone reproduced these associations (CDR r = 0.116 versus 0.121 with PGS616 added; RNFL r = −0.238 versus −0.204). The enrichment therefore reflects demographic risk. Quintile stratification revealed a stepwise decline in mean RNFL from 91.8 μm in the lowest-risk quintile to 83.6 μm in the highest-risk quintile, and a corresponding increase in mean CDR from 0.585 to 0.624 across the same range. IOP did not reach statistical significance (r = 0.034, p = 0.281).

To further characterize distributional differences by risk level, we compared the empirical cumulative distribution functions (ECDFs) of IOP, CDR, and RNFL thickness between individuals in the top and bottom 25% of MLP-predicted risk (Figure 4B). CDR showed a significant rightward shift in the high-risk group relative to the low-risk group (Kolmogorov–Smirnov [KS] p = 0.0121), indicating more pronounced optic nerve cupping. RNFL thickness showed an even stronger leftward shift in the high-risk group (KS p = 7×10□□), consistent with greater retinal nerve fiber thinning. IOP showed no significant distributional difference between risk groups (KS p = 0.135), consistent with the continuous correlation analysis.

### Phenotypic Asymmetry as a Clinically Meaningful Marker, Alone and with Genetic Risk Scores

In glaucoma diagnosis and risk stratification, asymmetry between the two eyes is considered an early sign of possible disease.^63,64^ In early-stage POAG, one eye may begin to show structural or functional changes while the other remains within normal limits.^64,65^ This asymmetry can manifest in IOP or the CDR, often before the disease becomes symmetric or clinically definitive. Moreover, since absolute values of these phenotypes were used to define training cases and controls (see *STAR Methods*), we derived inter-eye differences in IOP and CDR (ΔIOP, ΔCDR), defined as the absolute difference between the two eyes, to reduce feature overlap with the absolute-value thresholds used in case definition. Inter-eye RNFL difference (ΔRNFL) was not included because RNFL data were unavailable in the PMBB external validation cohort and had high rates of missingness (>30%) in the POAAGG suspect cohort. As illustrated in Figure 5A, inter-eye differences can reveal early glaucomatous changes even when absolute measurements fall within normal clinical ranges. These features carry strong clinical relevance and are especially valuable for detecting early or unilateral disease.

**Figure 5.**
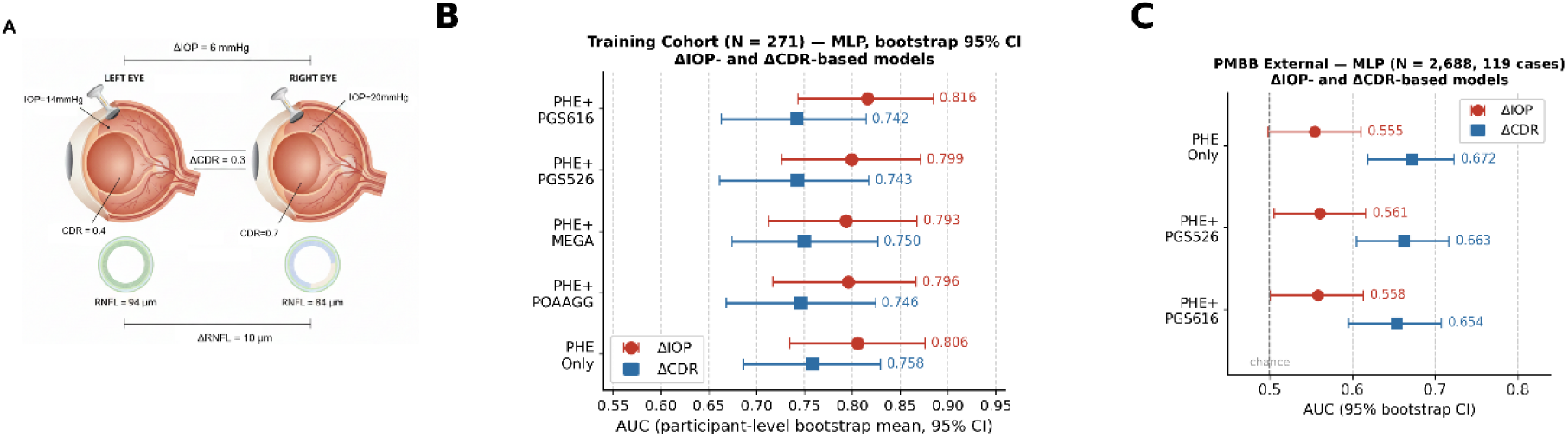
Inter-Eye Asymmetry Features for POAG Risk Prediction. (A) Schematic illustrating the derivation of inter-eye asymmetry features: ΔIOP (inter-eye intraocular pressure [IOP] difference) and ΔCDR (inter-eye cup-to-disc ratio [CDR] difference) are calculated as the absolute difference between right and left eye measurements. Asymmetry features were selected to reduce feature overlap with the absolute-value thresholds used in case definition. (B) Multilayer perceptron (MLP) area under the receiver operating characteristic curve (AUC; participant-level bootstrap estimate and 95% confidence interval [CI], B = 2,000; N = 271) for ΔIOP-based (red circles) and ΔCDR-based (blue squares) models, each trained with phenotype alone (PHE Only) or combined with one of four polygenic scores (PGS): POAAGG-PGS, MEGA-PGS, PGS526, or PGS616. ΔIOP models reached higher AUC than ΔCDR models in the training cohort across all feature combinations, although the confidence intervals overlap; adding PGS616 gave the highest nominal AUC for ΔIOP. (C) Penn Medicine BioBank (PMBB) external validation AUC (African ancestry; N = 2,688; 119 cases) for asymmetry model configurations with PHE Only, PHE+PGS526, and PHE+PGS616. The dashed vertical line indicates chance level (AUC = 0.5). Points are observed AUCs with 95% bootstrap CIs (B = 2,000). ΔCDR-based models (blue squares) generalize substantially better than ΔIOP-based models (red circles) externally; MLP models with and without PGS616 perform similarly (ΔCDR 0.672 and 0.654; ΔIOP 0.555 and 0.558); paired contrasts for all classifiers are in Table S18. ΔIOP models show lower external performance, possibly reflecting the greater clinical noise of IOP asymmetry in biobank settings; IOP-lowering medication use, which could attenuate true inter-eye IOP differences, could not be quantified in this dataset.

For each asymmetry phenotype, we trained disease-label classifiers using the asymmetry phenotype alone, referred to as the phenotype-only model, or the asymmetry phenotype plus one of four PGSs: POAAGG PGS, MEGA PGS, PGS526, or PGS616. ΔIOP- and ΔCDR-based models were trained separately to assess whether each PGS provided incremental information within each clinical domain. All four classifiers were evaluated using 5-fold × 20-repeat stratified cross-validation in the labeled POAAGG training cohort (N = 271). For the main figure, we present MLP results to maintain consistency with the primary clinical-enrichment, SHAP, and calibration analyses; results for all classifiers are provided in Table S18. External transportability of the asymmetry models (all four classifiers; Table S18) was assessed in the independent PMBB African ancestry cohort, which included POAG case-control labels and available IOP and CDR measurements.

Asymmetry features alone were moderately predictive (Figure 5B). For ΔIOP, the phenotype-only MLP model achieved a bootstrap AUC of 0.806 (participant-level bootstrap 95% CI: 0.734-0.876). Adding PGS616 gave the highest nominal value, 0.816 (0.743-0.885), but the paired increment, +0.010 (95% CI −0.037 to +0.043, p = 0.52; Table S19), was small. For ΔCDR, the phenotype-only model achieved AUC = 0.758 (participant-level bootstrap 95% CI: 0.686-0.830), and the four PGS gave similar values (paired increments −0.017 to −0.009, all p > 0.05; Table S19).

To assess external generalizability, we applied the asymmetry models to the PMBB African ancestry cohort. Unlike the 1,013-subject POAAGG suspect cohort used in the previous section (which lacked case/control labels), the PMBB cohort includes POAG case-control status, enabling direct AUC computation. We identified 2,688 PMBB participants of African ancestry aged ≥35 years with a baseline IOP or CDR measurement (119 POAG cases, 2,569 controls); the inter-eye difference could be computed for 2,657 participants for ΔIOP and 2,148 for ΔCDR, and missing differences were imputed with the training-cohort median. In PMBB external validation (Figure 5C), ΔCDR-based models generalized substantially better than ΔIOP-based models. For ΔCDR, the MLP phenotype-only model achieved an external AUC of 0.672 (95% CI: 0.619–0.723), and adding PGS616 gave 0.654 (95% CI: 0.595–0.708; paired ΔAUC −0.019, 95% CI −0.039 to +0.002, DeLong p = 0.08). For ΔIOP, the phenotype-only model yielded an external AUC of 0.555 (95% CI: 0.498–0.611), essentially unchanged by PGS616 (0.558, 95% CI: 0.501–0.613; paired ΔAUC +0.004, p = 0.68). For the primary MLP model, the asymmetry models therefore performed similarly with and without PGS616. The secondary classifiers were not uniform (Table S18): the random forest gained from either score with ΔCDR (PGS616 +0.054, p < 0.001; PGS526 +0.031, p = 0.02) and logistic regression gained slightly with ΔIOP (PGS616 +0.019, p = 0.03; PGS526 +0.018, p = 0.03), whereas the support vector machine and the MLP showed no significant change; these secondary comparisons are not adjusted for multiple testing.

## DISCUSSION

In this study, we evaluated four PGS for POAG and examined their performance when integrated with basic clinical information. Two scores captured genome-wide polygenic burden in African ancestry populations, while two were biologically curated and loci-based, drawing variants from six large multi-ancestry GWAS and applying effect size weights derived from African ancestry data. Across machine learning models the curated and genome-wide scores performed similarly, and the differences between them were small (Table S4), so the two approaches performed comparably in this setting. When combined with age and sex, the strongest configuration reached an internal AUC of up to 0.713 (MLP), and external performance remained similar to the demographic model (PMBB AFR, N = 9,084). Beyond case-control prediction, model-predicted risk aligned with established structural markers in an independent suspect cohort, an alignment that reflected its age and sex components.

First, our findings highlight the importance of developing risk models that are generalizable across genetically diverse populations. PGS trained in European ancestry populations frequently show reduced portability, especially in African ancestry groups with greater genetic diversity and distinct linkage disequilibrium structure. By constructing ancestry-matched genome-wide scores and curated loci-based scores weighted using African ancestry effect sizes, we evaluated whether population-aware PGS design improves prediction in an African ancestry population; in this cohort, performance was comparable to that of age and sex.

It is important to acknowledge the modest standalone predictive performance of PGS in this cohort. Genome-wide scores achieved near-chance discrimination (training AUC ≈ 0.46–0.54), while the best-performing standalone score, PGS526, reached a mean AUC of approximately 0.526 across classifiers, with an individual peak of 0.598 (RF). The confidence intervals of these values include 0.5 under participant-level bootstrap resampling (Table S4), so the scores are not intended as standalone diagnostic markers. This is expected given the complex, highly polygenic architecture of POAG and the substantial contribution of non-genetic factors (including IOP, age, and structural changes) to disease risk. The potential value of PGS therefore lies in a complementary contribution alongside clinical features; for the primary MLP model, the incremental AUC over age and sex was small in training (+0.001 by cross-validation) and in external validation (−0.001, DeLong p = 0.55).

We also observed a counterintuitive pattern in ancestry PC modeling: adding more PCs to the base model progressively reduced predictive performance, declining monotonically from Base+PC2 (mean AUC = 0.704) to Base+PC5 (0.688) to Base+PC10 (0.672). This suggests that beyond the first two PCs, additional ancestry axes introduce noise rather than informative signal in this well-characterized African ancestry cohort. In contrast, PGS616 achieved comparable training performance to PC2 alone (AUC = 0.700 vs. 0.704) while capturing partially distinct information: PC–PGS correlations were modest (max |r| = 0.36 for PGS616 with PC3, p < 0.001), and combining PC5 with PGS performed similarly to PGS alone (Base+PC5+PGS616: 0.688 vs. Base+PGS616: 0.700). This indicates that PGS616 is not simply a proxy for the ancestry axes. In ancestry PC-augmented models, PC1 was the dominant SHAP feature. SHAP describes the fitted model rather than the biological source of a signal, so this does not by itself show that these models learn population structure; together with the fact that the PC axes are not on a common basis across cohorts, however, it is consistent with their reduced performance in PMBB. In the PGS-augmented model, attributions were dominated by age and sex, with a smaller contribution from PGS616.

In the training cohort, the baseline demographic model achieved a mean cross-validated AUC of 0.683 (participant-level bootstrap 0.680, 95% CI: 0.605–0.748), consistent with the well-established role of age as a major risk factor for POAG. Adding PGS produced small, classifier-dependent increments over demographics in the training cohort, within sampling variability, with the strongest overall gain observed for Base+PGS616 (mean AUC = 0.700, +0.017 over baseline), and the best individual training performance achieved by LR and MLP with Base+PGS616 (AUC = 0.710–0.713). In the larger external biobank, the primary MLP model with Base+PGS616 reached 0.727 against a demographic baseline of 0.729 in PMBB (AFR, N = 9,084), and the cross-classifier average was 0.712 against 0.714 for age and sex alone. In conclusion, the polygenic scores evaluated here performed comparably to the demographic baseline. Our comparison of LR, RF, SVM, and MLP models highlights trade-offs between predictive performance and interpretability. All four classifiers achieved comparable performance using limited clinical and genetic features, with LR offering the added advantage of full interpretability, indicating that effective and transparent POAG risk prediction can be achieved without complex black-box models.

We also applied the models trained in the discovery cohort to an independent suspect cohort (N = 1,013). Using the MLP model trained on Base+PGS616, we generated individual risk scores for the 1,013 suspects and evaluated them against established clinical indicators (IOP, CDR, RNFL thickness). Higher MLP-predicted risk was significantly associated with larger CDR (r = 0.121, p < 0.001) and thinner RNFL (r = −0.204, p < 0.001), while IOP did not reach statistical significance. The absence of an IOP association is consistent with the known heterogeneity of IOP in early glaucoma suspect populations and the non-IOP-centric nature of the PGS features. Notably, CDR emerged as the most consistent correlate across model configurations, consistent with CDR being a more stable structural marker of cumulative glaucomatous damage than IOP, which is pharmacologically modifiable and may not fully reflect underlying disease biology. These results show that model-predicted risk tracks subclinical structural variation in an unlabeled cohort. Because this association was explained by age and sex, it reflects the demographic components of the model rather than a biological validation of the PGS.

The inter-eye asymmetry analyses provided a complementary assessment of clinically relevant POAG signal. Because ΔIOP and ΔCDR are derived from clinical domains related to POAG diagnosis, these results should be interpreted as biological consistency rather than fully independent validation. In the labeled POAAGG cohort, asymmetry features showed moderate discrimination of POAG status, and adding PGS616 changed them little (largest nominal increment +0.010, for ΔIOP with MLP, p = 0.52). In PMBB, ΔCDR-based models showed better external transportability than ΔIOP-based models, with similar MLP performance after adding PGS616 (ΔCDR 0.672 and 0.654, paired p = 0.08). Thus, inter-eye asymmetry appears to be a useful but context-dependent clinical signal. In the primary model its performance was similar with and without PGS616 in both cohorts, and some secondary classifiers showed small, classifier-dependent external gains.

Translation into clinical practice will require further validation. In this study the incremental contribution of the polygenic scores beyond age and sex was small in the training cohort and, for the primary model, in external validation, with similar case detection at fixed specificity. The analysis also provides a useful reference point: in a high-risk African ancestry population, age and sex form a strong baseline against which ancestry-matched polygenic scores can be benchmarked. Future scores can be evaluated against that baseline, in cohorts large enough to detect small increments.

In conclusion, this study provides a systematic evaluation of ancestry-matched genome-wide and curated PGS combined with basic clinical features for POAG risk identification in individuals of African ancestry. Age and sex formed a strong baseline, and adding PGS gave small, classifier-dependent increments; for the primary model, performance was comparable with and without PGS616 in the training cohort and in external replication in an independent biobank (PMBB, N = 9,084). Models trained on this framework stratify individuals along a disease gradient in an unlabeled suspect cohort, a stratification that reflects age and sex. Establishing the clinical value of ancestry-matched polygenic scores for POAG will benefit from larger cohorts, better-powered African ancestry discovery GWAS, and evaluation against a demographic baseline.

### Limitations of the study

This study has several limitations. The incremental predictive value of the PGS beyond age and sex was small, within sampling variability for every classifier in the training cohort under participant-level bootstrap resampling and for the primary MLP model in the external cohort (ΔAUC = −0.001, DeLong p = 0.55); externally it varied in sign across classifiers (from −0.025 for SVM to +0.011 for RF). Establishing clinical value will require larger, prospectively phenotyped cohorts. Age in PMBB was available only as age at the time of the data release, not at diagnosis or enrolment, so both the external age restriction and the age feature are approximations in that cohort. In addition, ancestry principal components were generated independently in the two cohorts, by different groups and by different procedures, so the axes are not on a common basis and PC-based features are not directly transferable across cohorts; this likely contributes to the reduced external performance of PC-augmented models. In addition, the clinical enrichment observed in the suspect cohort reflects age and sex: after adjustment for these covariates, the associations of the polygenic scores with cup-to-disc ratio, retinal nerve fiber layer thickness and intraocular pressure were small (Table S17). The training cohort (N = 271) and external validation cohort (PMBB, N = 9,084) were both recruited from a single academic medical center in Philadelphia, which may limit generalizability to other clinical settings and recruitment contexts. Although PGS were derived from multi-cohort African ancestry GWAS, prediction performance may still vary across African ancestry subgroups due to differences in allele frequencies and linkage disequilibrium patterns among continental African, African American, and admixed populations. Importantly, medication and treatment data, including IOP-lowering drug type, adherence, and duration, were not available as covariates; such treatment effects may confound observed relationships between clinical features and predicted risk, and future studies should account for this. Additionally, the binary case-control framework used for model training may oversimplify the continuum of POAG, and future models should explore probabilistic or multi-class labeling to better reflect real-world diagnostic uncertainty and support early detection.

Several modeling constraints also warrant consideration. The feature set was limited to available clinical variables; other glaucoma-relevant phenotypes such as visual field loss, optic disc morphology, and corneal hysteresis were excluded due to data sparsity or inconsistency, which may limit model comprehensiveness for atypical or advanced disease presentations. Learning curve analyses showed moderate train-to-validation gaps for the two-and three-feature models (0.065 and 0.083 at full training size) but a larger gap for Base+PC5+PGS616 (0.157), so larger cohorts would be needed to support higher-dimensional models. Finally, while complex classifiers such as MLP and RF introduce interpretability challenges, SHAP analysis showed that the primary model’s predictions are dominated by age and sex, consistent with established POAG risk epidemiology, whereas PC-augmented models were dominated by the first ancestry component.

## RESOURCE AVAILABILITY

### Lead contact

Further information and requests for resources, data, and code should be directed to and will be fulfilled by the lead contact, Joan M. O’Brien (joan.o’).

### Materials availability

This study did not generate new unique reagents.

### Data and code availability

- The de-identified genotype data from the POAAGG study are available under controlled access through dbGaP (accession phs001312).
- African ancestry external validation data from the Penn Medicine BioBank (PMBB) are available under controlled access through dbGaP (accession phs001453).
- Due to POAAGG subject privacy concerns and IRB regulations, identifiable data cannot be deposited in a public repository. However, de-identified datasets used for model training and evaluation are available from the lead contact upon request and pending data use agreement.
- Summary statistics and curated single nucleotide polymorphism (SNP) lists used to compute PGS are available in Table S2.
- Model performance tables, figure source data, and supplemental datasets supporting this manuscript are deposited at Mendeley Data: https://doi.org/10.17632/9rjn45y65c.2 (version 2, September 2026). Raw genotype data are not deposited there due to controlled-access restrictions (see dbGaP accessions above).
- All analysis scripts and notebooks are publicly available at: https://github.com/yzhu13/poag-pgs-ml (release v3-revision, https://github.com/yzhu13/poag-pgs-ml/releases/tag/v3-revision, corresponds to the analyses reported in this manuscript).

## Data Availability

The data underlying this study are not publicly available due to participant privacy and IRB restrictions. De-identified data may be made available from the corresponding author upon reasonable request and completion of a data use agreement. Code is available from the corresponding author upon request.

## ACKNOWLEDGMENTS

The POAAGG study was supported by the National Eye Institute, National Institutes of Health, through a Research Project Grant (R01EY023557) and a Vision Research Core Grant (2P30EY001583-51). Additional funding was provided by the F.M. Kirby Foundation, Research to Prevent Blindness, the University of Pennsylvania Hospital Board of Women Visitors, and the Paul and Evanina Bell Mackall Foundation Trust. Institutional support was received from the Department of Ophthalmology at the Perelman School of Medicine and the VA Hospital in Philadelphia, Pennsylvania. We acknowledge the Penn Medicine BioBank for providing data and thank the patient-participants of Penn Medicine who consented to participate in this research program. We would also like to thank the Penn Medicine BioBank team and Regeneron Genetics Center for providing genetic variant data for analysis. The PMBB is approved under IRB protocol #813913 and is supported by Perelman School of Medicine at University of Pennsylvania, a gift from the Smilow family, and the National Center for Advancing Translational Sciences of the National Institutes of Health under CTSA award number UL1TR001878. The funding sources had no role in the design and conduct of the study; collection, management, analysis, and interpretation of the data; preparation, review, or approval of the manuscript; or the decision to submit the manuscript for publication.

## AUTHOR CONTRIBUTIONS

Conceptualization: Yan Zhu, Shefali Setia-Verma, Joan M. O’Brien

Methodology: Yan Zhu, Aude Benigne Ikuzwe Sindikubwabo, Yuki Bradford, Lannawill Caruth

Software and Formal Analysis: Yan Zhu, Aude Benigne Ikuzwe Sindikubwabo, Yuki Bradford, Lannawill Caruth

Investigation: Yan Zhu, Rebecca Salowe, Laxmi Moksha, Vrathasha Vrathasha, Kenneth Pham, Roy Lee, Mina Halimitabrizi, Isabel Di Rosa, Leila Ghaffari, Jie He

Resources: Penn Medicine BioBank, Regeneron Genetics Center, Marylyn D. Ritchie, Shefali Setia-Verma, Joan M. O’Brien

Data Curation: Yan Zhu, Aude Benigne Ikuzwe Sindikubwabo, Penn Medicine BioBank, Regeneron Genetics Center

Writing – Original Draft: Yan Zhu, Rebecca Salowe

Writing – Review & Editing: Laxmi Moksha, Vrathasha Vrathasha, Kenneth Pham, Roy Lee, Mina Halimitabrizi, Isabel Di Rosa, Leila Ghaffari, Jie He, Joan M. O’Brien

Validation: Penn Medicine BioBank, Regeneron Genetics Center

Supervision: Marylyn D. Ritchie, Shefali Setia-Verma, Joan M. O’Brien

Funding Acquisition: Joan M. O’Brien

All authors have read and agreed to the published version of the manuscript.

## SUPPLEMENTAL INFORMATION

Document S1. Supplemental Figures S1–S7, Related to STAR Methods.

Figure S1. Full Circos layout of genetic variant distribution across autosomes.

Figure S2A. Pearson correlations between ancestry principal components and polygenic risk scores.

Figure S2B. Ancestry PC-augmented versus PGS-augmented model performance with participant-level bootstrap confidence intervals.

Figure S3. SHAP feature importance, calibration curves, and Brier score assessments.

Figure S4. Learning curves across four machine learning classifiers.

Figure S5. Sex-stratified AUC analysis with participant-level bootstrap confidence intervals.

Figure S6. Incremental AUC (ΔAUC) of PGS616 over the age + sex baseline, related to Figure 3.

Figure S7A. Predicted risk correlations with clinical outcomes across all classifiers and feature sets.

Figure S7B. MLP-predicted POAG risk vs. individual clinical outcomes in the suspect cohort.

Table S1. Demographic and clinical characteristics of the training, suspect and external validation cohorts, related to Figure 1.

Table S2. SNP lists and African ancestry effect sizes for curated PGS526 and PGS616, related to Figure 2.

Table S3. Construction provenance and variant accounting for PGS616 and PGS526, related to STAR Methods.

Table S4. Standalone PGS discrimination and comparison of curated versus genome-wide scores, related to Figure 2.

Table S5. Model performance metrics across feature sets in the training cohort (5×20 CV), related to Figure 2 and Figure 3.

Table S6. Same-classifier AUC comparison (Base vs. Base+PGS), related to Figure 3.

Table S7. Incremental AUC (ΔAUC) with confidence intervals and significance, related to Figure 3.

Table S8. Participant-level bootstrap AUC and incremental ΔAUC, training cohort, related to Figure 3.

Table S9. PC–PGS correlation coefficients and p-values, related to Figure S2.

Table S10. PGS residualized on ancestry principal components, related to Figure 3.

Table S11. SHAP feature importance values for MLP configurations, related to Figure S3.

Table S12. Calibration metrics and Brier scores, related to Figure S3.

Table S13. Learning curve data and train-to-CV gaps, related to Figure S4.

Table S14. Sex-stratified performance metrics, related to Figure S5.

Table S15. PMBB external validation performance metrics, related to Figure 3.

Table S16. Statistical versus clinical significance of the PGS616 increment, related to Figure 3.

Table S17. Suspect cohort: PGS association with ocular phenotypes adjusted for age and sex, related to Figure 4.

Table S18. Asymmetry-based model performance metrics across classifiers, related to Figure 5.

Table S19. Participant-level bootstrap intervals for the sex-stratified, inter-eye asymmetry, and PGS-residualization analyses, related to Tables S10, S14 and S18.

Table S20. Data-leakage safeguards by pipeline stage, related to STAR Methods.

## STAR⍰METHODS

### KEY RESOURCES TABLE

| REAGENT OR RESOURCE | SOURCE | IDENTIFIER |
| --- | --- | --- |
| Deposited data |  |  |
| Genotype data for ADAGES participants | dbGaP | Accession phs001673 |
| GGLAD POAG GWAS summary statistics | Hauser et al.<sup>66</sup> | https://s3-ap-southeast-1.amazonaws.com/gis-hg4-poag/poag_africa_4_collection_logistic.meta.sorted_N4.tsv.zip |
| Genotype data for POAAGG participants | dbGaP | Accession phs001312 |
| PMBB | Penn Medicine BioBank | https://pmbb.med.upenn.edu/ (PMID: 36556195) |
| Software and algorithms |  |  |
| Python 3.13 (3.11 previously; identical results) | Python Software Foundation | RRID:SCR_008394 |
| R 4.2.2 | https://www.r-project.org | https://cloud.r-project.org/bin/windows/base |
| Scikit-learn 1.7.2 (1.3.0 previously; identical results) | scikit-learn developers<sup>67</sup> | RRID:SCR_002577 |
| TOPMED Imputation Reference Panel | Das et al.<sup>68</sup> | https://imputation.biodatacatalyst.nhlbi.nih.gov/#! |
| EIGENSOFT package | Galinsky et al.<sup>69,70</sup> | http://www.hsph.harvard.edu/alkes-price/software |
| SAIGE version 1.1 software | Mbatchou et al.<sup>71</sup> | https://saigegit.github.io/SAIGE-doc/ |
| Matplotlib / Seaborn | Python | N/A |
| PRS-CS | https://github.com/getian107/PRScs | N/A |
| PLINK v1.9 / v2.0 | Chang et al.<sup>72,73</sup> | https://www.cog-genomics.org/plink/ |
| Other |  |  |
| Google Colab environment | https://colab.research.google.com | N/A |
| GitHub repository | https://github.com/yzhu13/poag-pgs-ml | N/A |

## EXPERIMENTAL MODEL AND STUDY PARTICIPANT DETAILS

### Human Study Participants

Three cohorts contributed to this study: the POAAGG study cohort (PGS generation and ML model training/validation), the MEGA dataset (PGS generation), and the PMBB cohort (external ML validation).

**(1) POAAGG Study**

The POAAGG study cohort consists of self-identified Blacks (African Americans, African descent, or African Caribbean), 35 years or older, identified from the Philadelphia region. Each subject was classified as a case, control, or suspect by a glaucoma specialist or ophthalmologist, based on previously detailed criteria.^74^ Subjects underwent a comprehensive onsite interview and examination, including the collection of demographic information and comprehensive ocular phenotypes. All subjects signed an informed consent form and provided a genomic DNA sample. ^9,60,75^ The study was approved by the Institutional Review Board at the University of Pennsylvania and was conducted in accordance with the tenets of the Declaration of Helsinki.

For the current analysis, a total of 8,315 POAAGG subjects (including 2,723 cases, 4,579 controls, and 1,013 suspects) were included. All subjects had available genotype data, demographic information (age, gender), and at least one ocular phenotype (IOP, vertical CDR, and RNFL thickness). This comprised:

- **POAAGG PGS Generation cohort** (N=7,031, with 2,595 cases and 4,436 controls): This cohort was used in the generation of the POAAGG PGS, described in the below section.
- **Training cohort (N=271, with 128 cases and 143 controls):** This cohort consists of cases and controls not included in the generation of the PGS scores. To ensure that subjects correctly fit their classification of case or control, we carefully reviewed their phenotypes. 99 clean cases met the following phenotypes in both eyes: IOP ≥ 22.1 mmHg, CDR ≥ 0.7, and RNFL ≤ 80 µm. 97 clean controls had normal phenotypes for both eyes, including: IOP between 10–16 mmHg, CDR ≤ 0.5, and RNFL ≥ 82 µm. We acknowledge that these thresholds may enrich the training set for phenotypically extreme individuals, potentially introducing spectrum bias and yielding optimistic performance estimates relative to more heterogeneous clinical populations. To partially mitigate this, the remaining 75 subjects were drawn from recently accrued, real-world clinical POAG cases and controls within the same healthcare system without strict phenotypic filtering, introducing greater clinical heterogeneity into the training data. External validation in the independent PMBB cohort (N=9,084) provides a separate assessment of model generalizability beyond the training distribution (Table S15).
- **Suspect cohort** (N=1,013): This cohort was used to assess model generalizability and perform clinical enrichment analyses.^60^ The individuals in the suspect cohort were not included in PGS generation and were not used in model training.
- **(2) MEGA Dataset**

The MEGA dataset was used to generate the MEGA PGS, as described previously.^14^ This cohort consists of 11,275 subjects (6,003 cases and 5,272 controls) from three African ancestry glaucoma cohorts: (1) the African Descent and Glaucoma Evaluation Study (ADAGES, n=1,999), (2) Genetics of Glaucoma in People of African Descent (GGLAD, n=2,952)^66^, and (3) POAAGG study (n=6,324). The POAAGG component of the MEGA dataset (n=6,324) overlaps with the POAAGG PGS generation cohort (n=7,031), as both draw from the broader POAAGG study population; however, these cohorts were used solely for summary-statistic-level GWAS and PGS weight derivation. Critically, the 271-subject ML training cohort was excluded from both the POAAGG PGS generation cohort and the MEGA dataset’s POAAGG component, ensuring no individual-level data leakage between PGS construction and model training.

**(3) PMBB Cohort**

PMBB is a large, health system-based biorepository at the University of Pennsylvania.^59^ Approximately 350,000 participants have enrolled to date, of whom 57,170 have genome-wide genotype data imputed to the TOPMed reference panel.^59^ PMBB also integrates comprehensive electronic health record (EHR) information, including diagnostic codes, laboratory values, imaging results, medication histories, and selected lifestyle factors.

We included PMBB participants with genetically inferred African ancestry and aged ≥35 years at the time of the data release, the only age field available in the phenotype file. Ancestry was determined using principal component analysis (PCA). Principal components were generated independently for the two cohorts by different groups and by different procedures. For POAAGG, genotypes were merged with samples from the 1000 Genomes Project reference panel and principal components were computed on the combined dataset by eigen-decomposition using EIGENSOFT v8.0,^69,70^ after filtering to minor allele frequency ≥ 0.01 and ≥ 99% call rate and pruning for linkage disequilibrium (--indep-pairwise 100 10 0.01); twenty components were retained. For PMBB, principal components were produced by the Penn Medicine BioBank Informatics group as part of the standard PMBB data release. Because the two sets of components derive from different procedures applied to different sample sets, the resulting axes are not on a common basis and are not comparable across cohorts; accordingly, our primary model (age, sex, and PGS616) does not include PCs, and PC-augmented models were evaluated only in secondary analyses. Related individuals were retained. POAG cases were defined using harmonized ICD-based criteria across cohorts. Specifically, cases included individuals with ICD-9 codes 365.10 or 365.11, or any ICD-10 H40.11 variant (primary open-angle glaucoma). Individuals with ICD-10 codes H40.01* (borderline findings) and H42* (glaucoma in diseases classified elsewhere) were excluded to remove secondary glaucoma due to trauma, inflammation, drug effects, or other underlying conditions. Controls were defined as individuals with no evidence of glaucoma, including no ICD-9 codes beginning with 365 and no ICD-10 codes within the H40 or H42 ranges.

## METHOD DETAILS

### 1. Data Preprocessing and Cohort Selection

This study used two distinct categories of data sources with non-overlapping roles: (i) large-scale genetic datasets for PGS derivation and (ii) a deeply phenotyped clinical cohort for machine learning model development and validation.

### PGS derivation datasets

PGS were derived using GWAS summary statistics and individual-level data from African ancestry subjects in the POAAGG study and the MEGA cohort, as described above. All MEGA data were imputed to the TOPMed reference panel,^68,76^ and association testing accounted for population structure and relatedness^71^. These datasets were used exclusively for PGS construction and weighting and did not contribute to machine learning model training or testing.

### Machine learning training and validation cohorts

All machine learning model training and internal evaluation were performed within the POAAGG cohort. A curated case-control subset (N=271) was used for model training and 5-fold × 20-repeat stratified cross-validation (5×20 CV; 100 total model fits), while an independent suspect cohort (N=1,013) was used for biological validation and clinical enrichment analyses. External validation was conducted in the independent PMBB cohort (N=9,084 African ancestry participants). No subjects used for PGS derivation were included in model training or testing, ensuring strict separation between genetic discovery and predictive modeling.

Demographic features were numerically encoded (male = 1, female = 0). Continuous clinical traits, including IOP, vertical CDR, and RNFL thickness, were standardized using z-score transformation. Missing values were imputed with medians estimated in the training data (within each training partition during cross-validation and bootstrap resampling) and applied unchanged to held-out and external data. All preprocessing steps, including imputation and scaling, were implemented within scikit-learn pipelines to ensure consistent application across cross-validation iterations and prevent data leakage.^67^

### 2. PGS Construction

Four PGS were constructed using two complementary strategies (genome-wide Bayesian shrinkage and curated locus-based selection), all computed using African ancestry-specific effect sizes. The four scores are:(1) The POAAGG PGS, based solely on GWAS summary statistics from the POAAGG cohort (N = 7,031),^9,77^ and (2) The MEGA PGS, based on a larger African ancestry GWAS mega-analysis (MEGA, N = 11,275) combining three cohorts: POAAGG, ADAGES, and GGLAD.^14,15^ The second strategy focused on biologically curated, loci-based scores. We systematically extracted genome-wide significant SNPs from six large glaucoma GWAS across multiple ancestries: Craig et al. (2020),^45^ who identified 114 POAG-associated loci in a multi-ancestry meta-analysis; Gharahkhani et al. (2021),^46^ who identified 127 loci associated with POAG and IOP across European and Asian ancestries; Han et al. (2023),^49^ who reported 312 loci from a multi-ancestry meta-analysis and 263 loci from a European ancestry multitrait analysis; Lo Faro et al. (2024),^47^ who reported 62 POAG-associated loci from the Global Biobank Meta-analysis Initiative (GBMI); Chang-Wolf et al. (2025),^48^ who reported POAG-associated loci in European ancestry populations; and Verma et al. (2024),^14^ who identified 46 POAG risk loci in African ancestry individuals from the POAAGG cohort. All loci originally reported in GRCh37/hg19 coordinates were converted to GRCh38/hg38 using UCSC liftOver (chain file hg19ToHg38). SNPs were then cross-referenced against the MEGA GWAS summary statistics to retain only variants with available African ancestry effect size estimates, yielding an initial pool of 572 unique SNPs from the five non-Verma discovery datasets (see Variant accounting below), plus 46 from Verma et al. These variants were then weighted using effect sizes derived from the African ancestry MEGA GWAS (PGS616) or the POAAGG quantitative multi-trait analysis of GWAS (MTAG) (PGS526), and filtered based on PLINK 2.0 scoring compatibility, resulting in two distinct curated scores: (3) PGS526, the 526 variants carrying a usable MTAG weight, whose weights emphasize IOP and optic nerve structure (CDR, RNFL), and (4) PGS616, the 616 variants carrying a usable MEGA case–control weight. The curated SNPs are lead variants from discovery GWAS that each applied their own linkage disequilibrium (LD) clumping; no additional LD pruning was applied to the combined panel, so residual LD between variants drawn from different studies cannot be excluded. SNP selection, weighting method, filtering, LD pruning, and normalization procedures for each score are detailed below. A complete list of SNPs and weights for PGS526 and PGS616, including African ancestry MEGA GWAS p-values and study sources, is provided (Table S2). Standard errors were not available for all source variants; p-values and effect-size sources are provided where available.

Note on African-specific effect sizes and MEGA significance: PGS616 and PGS526 use African ancestry-specific effect sizes from the MEGA GWAS and MTAG, respectively. Because MEGA (N=11,275) is substantially smaller than the multi-ancestry discovery cohorts, many curated SNPs do not reach genome-wide significance (p < 5×10⁻ □) in the MEGA GWAS itself (45/616 and 40/526 SNPs reach GWS in MEGA, respectively); their MEGA p-values and effect sizes are provided in Table S2 for transparency. The inclusion of these loci is justified by their genome-wide significant discovery in the original multi-ancestry GWAS and by the use of directional MEGA African ancestry weights, which provide ancestry-matched calibration even for loci with limited power in MEGA alone.

1. POAAGG PGS: The POAAGG PGS is a genome-wide polygenic score derived from GWAS summary statistics generated within the POAAGG cohort (N = 7,031), which includes African ancestry subjects. We used the PRS-CS framework, a Bayesian method that applies continuous shrinkage priors to SNP effect sizes while accounting for LD.^44^ Effect size modeling was performed using the African ancestry reference panel from the 1000 Genomes Project, with default parameters (φ = auto, 1,000 burn-in and 2,000 MCMC iterations).^14,26^ Ambiguous SNPs (A/T or G/C) were removed prior to scoring.^78^ Genome-wide scores were computed using the weighted sum of alleles in PLINK 2.0 and then normalized with the rank-based inverse normal transformation described for PGS526 below.^72,73^ This score reflects cohort-specific polygenic risk and serves as a comparator for broader African ancestry models.
2. MEGA PGS: The MEGA PGS was constructed using the same methodology as the POAAGG PGS but applied to the mega-analysis of African ancestry individuals (N=11,275) drawn from three large glaucoma cohorts (ADAGES, GGLAD, and POAAGG). By aggregating individual-level data across multiple cohorts, this score captures a more comprehensive and generalizable estimate of POAG polygenic burden in African ancestry populations. The POAAGG component of this mega-analysis (n=6,324) overlaps with the POAAGG PGS generation cohort, as both draw from the broader POAAGG study population; the 271-subject ML training cohort was excluded from this POAAGG component, ensuring no data leakage between MEGA PGS construction and model training. As with the POAAGG PGS, we used PRS-CS with the African ancestry LD reference panel and computed weighted allele sums in PLINK 2.0,^72,73^ followed by the same rank-based inverse normal transformation.
3. PGS526: PGS526 is a biologically curated, locus-based score composed of the 526 curated SNPs (after liftover and MEGA overlap filtering as described above) that carry a usable effect-size estimate in the POAAGG quantitative multi-trait analysis (MTAG) described below; it is therefore a subset of the PGS616 panel, and the two scores differ principally in their weights (Table S3). SNPs were drawn from six large multi-ancestry glaucoma GWAS (Craig et al.,^45^ Gharahkhani et al.,^46^ Han et al.,^49^ Lo Faro et al.,^47^ Chang-Wolf et al.,^48^ Verma et al.^14^), harmonized to the hg38 reference genome (using UCSC liftOver, chain file hg19ToHg38), and filtered to retain variants present in the MEGA summary statistics. Effect sizes for PGS616 were derived from the MEGA African ancestry case/control GWAS. Effect sizes for PGS526 were derived from the POAAGG quantitative multi-trait meta-analysis (MTAG), which integrates GWAS signal from IOP, CDR, and RNFL endophenotypes alongside POAG case/control status, yielding ancestry-matched weights optimized for endophenotype-related risk.^14^ Final scores were computed in PLINK 2.0,^72,73^ normalized using a rank-based inverse normal transformation, qnorm((rank(x) - 0.5)/n), computed within each cohort in which the score was calculated: across the 1,284 POAAGG participants of the training and suspect cohorts together, and across all 56,357 PMBB participants with computed scores, before the ancestry and age restrictions were applied. The transform uses no outcome labels and carries no parameters between POAAGG and PMBB, so no information passes from the training cohort to the external validation cohort through the standardization step; within POAAGG the training and suspect cohorts share one transformation. Scores are consequently cohort-relative.^72,73^ PGS526 emphasizes mechanistic genetic risk relevant to measurable physiological traits underlying POAG.
4. PGS616: PGS616 is a complementary, selected locus-based PGS designed to capture broader POAG disease risk by integrating 616 SNPs (the final count after liftover and MEGA overlap filtering) derived from genome-wide significant POAG-associated loci across the same six GWAS used for PGS526. PGS616 and PGS526 share the same curated panel and differ in their weights: PGS616 uses MEGA case–control effect sizes for POAG disease status, and PGS526 uses POAAGG MTAG effect sizes that emphasize the endophenotypes.^14^ The additional 90 SNPs in PGS616 relative to PGS526 reflect disease-status loci that are included based on MEGA case/control GWAS evidence but lack a usable effect-size estimate in the POAAGG MTAG used for PGS526. All variants were subjected to the same filtering and weighting pipeline as PGS526, with African ancestry-specific effect sizes drawn from MEGA summary statistics. Like the other scores, PGS616 was computed using PLINK 2.0 and normalized using the same rank-based inverse normal transformation and standardization procedure as PGS526.^72,73^ Together with PGS526, this score represents a curated, interpretable risk model that highlights genetic signals with strong disease-specific relevance in African ancestry populations.

These four scores represent complementary genetic risk models that balance breadth, biological specificity, and ancestry-matched calibration, forming the foundation for downstream machine learning and clinical enrichment analyses.

### Variant accounting

Genome-wide significant loci were extracted from multiple discovery datasets, giving 878 loci in total (Craig 114, Lo Faro 62, Gharahkhani 127, Han multi-ancestry 312, Han European ancestry 263). Coordinates reported in GRCh37 were converted to GRCh38 with UCSC liftOver (chain hg19ToHg38); the Lo Faro table was already in GRCh38. Variants were retained if present in the MEGA African ancestry summary statistics, leaving 790 loci, deduplicated across sources to 572 unique variants, and combined with the 46 African ancestry POAG loci from Verma et al. to give 618. Of these, 616 carried usable MEGA case–control effect sizes (PGS616) and 526 carried usable POAAGG MTAG effect sizes (PGS526). The complete accounting, with software versions and commands, is given in Table S3.

### Genotype-level quality control and scoring

Extraction of the curated variants from the POAAGG genotype panel by position returned 622 genotype records (a record count: a multi-allelic site contributes one record per alternate allele); 54 records were removed at genotype quality control, chiefly for missingness of 15% or more (overall genotyping rate 0.931), leaving 568. Scores were computed in PLINK v2.00a6LM with --score and the scoresums modifier, which mean-imputes residual missing dosages to the cohort allele frequency. PGS616 was computed from 568 scored variants and PGS526 from 486; the scores are named for the number of variants carrying usable weights rather than the number present in this genotype panel. Genotype quality control and file preparation used PLINK v1.90p. In PMBB, 539 PGS616 and 460 PGS526 variants were scored, of which 525 and 448 were also scored in POAAGG; rescoring both cohorts on these shared variants left the primary external result unchanged (MLP ΔAUC for PGS616 +0.002, DeLong p = 0.65; Table S3).

### Linkage disequilibrium

No LD pruning was applied beyond that performed by the discovery GWAS. The curated variants are lead SNPs from studies that each applied their own LD-based clumping; no additional pruning step was performed at any stage and no joint LD check was made across studies, so some residual LD between variants from different sources may remain.

### 3. Machine Learning Models: LR, RF, MLP, and SVM

We applied four ML classifiers to each feature set: LR, RF, MLP, and SVM, implemented using the scikit-learn library (v1.7.2).^67^ Each classifier was trained to distinguish POAG cases from controls based on different combinations of demographic, clinical, and genetic features.

- LR was implemented with class_weight=’balanced’ and max_iter=1000.^50,51^
- RF was configured with 200 decision trees, max_depth=5, and class_weight=’balanced’ to correct for label imbalance.
- MLP employed a single hidden layer with 32 neurons and max_iter=1000; early stopping was disabled (early_stopping=False) to ensure stable convergence across all cross-validation folds.
- SVM was used with an RBF kernel (C = 1.0, gamma =’scale’), class_weight=’balanced’, and probability=True to support AUC calculation. For LR, the regularization strength was C = 1.0 (L2 penalty, default). No hyperparameter search or grid search was performed; all parameters were fixed a priori based on published recommendations for imbalanced binary classification. All classifiers were implemented in scikit-learn (v1.7.2) with a preprocessing pipeline of median imputation followed by z-score standardization.^67^ Performance variability across the 100 CV iterations (5 folds x 20 repeats) is reported as mean +/- SD AUC and is descriptive only. Because the 100 fold-level estimates are not independent, confidence intervals and significance tests for the training cohort are derived from a participant-level bootstrap instead (see Uncertainty estimation in the training cohort).

Model performance for standalone PGS evaluation (Figure 2C) was assessed using 5-fold × 20-repeat stratified cross-validation (5×20 CV), yielding 100 total model fits per configuration.^79,80^ In each repetition, the training cohort (N□=□271) was partitioned into 5 stratified folds preserving the case-control ratio; models were trained on 4 folds and evaluated on the held-out fold, repeated 20 times with different random seeds to ensure robust estimation.^79^ The training pipeline included median imputation for missing values (SimpleImputer), z-score standardization of continuous features (StandardScaler), and model fitting.^81,82^

Three families of models were fitted: standalone PGS models (each score alone; Figure 2C), the case–control feature sets listed below, and the inter-eye asymmetry models described in section 5. In the case–control feature sets each PGS was incorporated as a continuous variable alongside age and sex. To systematically evaluate the role of ancestry PCs, we constructed twelve primary feature sets:

- (1) Age only
- (2) Sex only
- (3) Base = age + sex
- (4) Base + PC2 (age + sex + PC1–PC2)
- (5) Base + PC5 (age + sex + PC1–PC5)
- (6) Base + PC10 (age + sex + PC1–PC10)
- (7) Base + POAAGG PGS
- (8) Base + MEGA PGS
- (9) Base + PGS526
- (10) Base + PGS616
- (11) Base + PC5 + PGS526
- (12) Base + PC5 + PGS616

Items (7)–(10) follow the same structure as the original five-set design; items (4)–(6) were added to evaluate PC confounding; items (11)–(12) test whether PCs and PGS provide additive information. This structured design enabled systematic assessment of the individual and combined contributions of demographic, ancestry, and genetic features to glaucoma risk prediction. External validation in the PMBB cohort was performed for six of the twelve feature sets (Base, Base+PGS526, Base+PGS616, Base+PC5, Base+PC5+PGS526, Base+PC5+PGS616), using matched covariates (age, sex, PC1–PC5, and PGS). Two further feature sets, Base+PC20 and Base+PC20+PGS616, were evaluated in the training cohort as sensitivity analyses of the number of retained components (Table S5); Base+PC20 is also included in the participant-level bootstrap (Table S8). Full feature definitions and modeling pipelines are publicly available at: https://github.com/yzhu13/poag-pgs-ml.

Performance metrics, including AUC, accuracy, precision, sensitivity, specificity, and F1-score, were computed on the held-out fold for each cross-validation split. Final reported values represent the mean and standard deviation across the 100 cross-validation iterations (5 folds × 20 repeats).^80^ This cross-validation strategy was applied consistently across all four classifiers (LR, RF, MLP, SVM) and all twelve feature set configurations to ensure fair model comparison and robust performance estimation. For PMBB external validation, models were trained on the full POAAGG training set (N = 271) and applied directly to the PMBB African ancestry subset (N = 9,084); bootstrap 95% CIs (B = 2,000 participant-level replicates) were computed for PMBB AUC estimates.

### 4. Testing Cohort Analysis and Clinical Enrichment in Suspect Population

To evaluate the biological relevance and generalizability of PGS-informed models in the absence of definitive diagnostic labels, trained ML models were applied without retraining to an independent suspect cohort (N = 1,013). Models were trained exclusively in the labeled training cohort (N = 271) and then used to generate continuous, individual-level predicted glaucoma risk scores for all suspects.

For primary visualization, we focused on the MLP model with the Base+PGS616 feature set to maintain consistency with the primary prediction, SHAP, and calibration analyses; full classifier-by-feature-set results are shown in Figure S7A–B. Predicted risk scores were retained as continuous individual level outputs for statistical inference. Associations between predicted risk and quantitative ocular phenotypes (IOP, CDR, RNFL thickness) were evaluated using Pearson correlation across all individuals with available phenotype data. For visualization, predicted risk scores were stratified into quintiles (1–5), mean phenotype values within each quintile were plotted with ± SEM, and an OLS trend line was fit to the five quintile means to illustrate the directional gradient.

For visualization of enrichment patterns, predicted risk scores were stratified into quintiles (1–5), and mean phenotype values within each quintile were plotted with corresponding standard errors (± SEM). Because the suspect cohort lacks definitive diagnostic labels, classification metrics such as AUC could not be computed; therefore, model evaluation focused on clinical enrichment analyses using continuous statistical testing complemented by stratified visualization.

In addition, ECDFs were used to compare phenotype distributions between individuals in the top and bottom quartiles of predicted risk. Differences between distributions were evaluated using the Kolmogorov–Smirnov test.

This enrichment-based validation framework enables assessment of biological plausibility and disease relevance in unlabeled populations and is consistent with prior approaches for evaluating risk prediction models in real-world clinical cohorts.

### 5. Asymmetry Analysis and Biological Validation

To evaluate inter-eye asymmetry as an independent and clinically meaningful disease signal while avoiding circularity from cohort definition criteria, we derived inter-eye difference features for ΔIOP and ΔCDR.^63,65,83^ ΔRNFL was excluded from asymmetry features to ensure consistency across all three cohorts (POAAGG training, suspect, and PMBB external), as RNFL measurements were unavailable in the PMBB dataset. Each asymmetry feature was calculated as the absolute difference between the right and left eyes (|Right − Left|).^83^

ΔIOP and ΔCDR were evaluated independently as asymmetry features, each alone and in combination with each of the four PGS scores, under the same 5×20 CV framework and classifier configurations described above (LR, RF, MLP, SVM; N = 271 training cohort). MLP results are shown in Figure 5B for consistency with the primary model used in the suspect-cohort, SHAP and calibration analyses; results for LR, SVM, and RF are provided in Table S18, with participant-level bootstrap intervals in Table S19. External validation was performed in the PMBB African ancestry cohort for the phenotype-only (PHE-only), PHE+PGS526, and PHE+PGS616 configurations (Figure 5C), where matched IOP and CDR phenotype data were available; bootstrap 95% CIs (B = 2,000 participant-level replicates) were computed for PMBB AUC estimates.

### 6. Software and Reproducibility

All data preprocessing, feature engineering, and ML modeling were performed in Python (version 3.13 for the analyses reported in this revision; version 3.11 for the previously reported results, which reproduce exactly under both)^84^ using open-source libraries, including scikit-learn (version 1.7.2 for the analyses reported in this revision; version 1.3.0 previously, with identical results)^67^ for model implementation, pandas and numpy for data manipulation, matplotlib for visualization, SciPy for statistical analysis, and joblib for parallel execution of the bootstrap resampling. Genetic data processing, including risk score calculation and dataset merging, was conducted using PLINK v2.0^73^ and PRS-CS^44^, with ancestry-specific LD panels used to infer SNP weights for PGS.

To address potential data leakage, we note that the 271-sample training cohort is a distinct subset of the 8,315-subject POAAGG study and is entirely separate from both the PGS generation cohort and the PMBB cohort used for external validation. Specifically, the POAAGG PGS was derived from GWAS summary statistics generated using a dedicated PGS generation cohort of 7,031 POAAGG subjects; the 271 training cohort subjects were explicitly excluded from this GWAS computation and therefore had no influence on the PGS effect size estimates used in model training. The PMBB cohort is an independent biobank with no overlap with any POAAGG subjects. The curated loci (PGS526/PGS616) were selected based on independent GWAS evidence from separate cohorts (Craig et al., Han et al.), with no optimization against the 271-sample training set.

The original analysis scripts were developed and executed within Google Colaboratory (Colab)^85^, facilitating a cloud-based computational environment with consistent software dependencies across users; the analyses repeated for this revision were run locally in Python, with the versions listed in the key resources table. The project followed reproducible research practices, with each model training and evaluation task encapsulated in modular code blocks with clearly defined input and output interfaces. Outputs for each model–feature set combination were stored as labeled CSV and Excel files, with naming conventions reflecting the model type (e.g., LR, RF, MLP, SVM), feature set, and performance metrics.

To promote transparency and facilitate replication, all analysis scripts and notebooks, including those used for 5-fold × 20-repeat stratified cross-validation and visualization generation, have been deposited in a GitHub repository and are available publicly, as described in the Data and Code Availability section.

All visualizations presented in the main text were generated directly from model outputs using standardized plotting functions that support error bar overlays and stratified grouping. These functions are also included in the repository and can be reused or adapted by other researchers. Together, this combination of modular scripting, version controlled code, and cloud based execution supports full reproducibility of the AI modeling pipeline and facilitates broader adoption of these tools for glaucoma risk prediction.

### 7. Ancestry Principal Components: Construction, Selection, and Confounding Assessment

We evaluated PC-augmented feature sets (Base+PC2, Base+PC5, Base+PC10, Base+PC20) under rigorous 5×20 CV (RepeatedStratifiedKFold, n_splits=5, n_repeats=20, random_state=42). For Base+PC20, the twenty ancestry principal components described above (columns PC1–PC20 in the training data file) were added to the Base feature set. The principal components were computed once, on a set of 1,285 POAAGG participants that includes both the 271-participant modeling cohort and the 1,013-participant suspect cohort, so those participants contributed to the eigen-decomposition. The decomposition is unsupervised and used no case-control labels, so no outcome information entered the components; the components are nevertheless not independent of the modeling sample, and were not recomputed within cross-validation folds or bootstrap replicates. Internal performance of PC-augmented models may therefore be slightly optimistic relative to their performance on genuinely external data, which is a further reason, alongside the difference in construction between cohorts, for the reduced external performance of these models. PC-augmented feature sets are secondary analyses throughout and the primary model contains no principal components. All classifier pipelines (LR, SVM, RF, MLP) were evaluated. PC-augmented results were compared to PGS-augmented models and to the externally validated performance in PMBB. Results are presented in Figure S2, Table S5 and Table S9.

### Components retained

Twenty principal components are available for the POAAGG cohort. Rather than fixing a single number a priori, we treated the number retained as an explicit modeling choice and evaluated four nested feature sets, Base+PC2, Base+PC5, Base+PC10 and Base+PC20, under identical cross-validation and bootstrap procedures, so that the effect of retaining more ancestry axes could be read directly from model performance.

### Incorporation into models

Principal components enter as continuous covariates alongside age and sex and are standardized within the modeling pipeline. They are not used to stratify, match, or weight participants. The primary model (age, sex and PGS616) contains no principal components; PC-augmented feature sets are secondary analyses reported for the confounding assessment described below.

### Assessment of ancestry confounding

Three analyses address whether the polygenic scores act as proxies for ancestry. First, Pearson correlations between PC1–PC5 and each PGS quantify their linear overlap (Table S9, Figure S2A). Second, each PGS was regressed on PC1–PC5 within cohort and the residual re-entered the models in place of the raw score, quantifying how much of the score’s contribution survives removal of its ancestry-correlated component (Table S10; bootstrap intervals in Table S19). Third, PC-augmented and PGS-augmented feature sets were compared directly, internally and externally (Figure 3, Figure S2B).

### 8. SHAP Feature Importance Analysis

To provide model-agnostic feature importance estimates, SHAP analysis was performed using the KernelExplainer from the SHAP library (version ≥0.40).^61^ The MLP classifier was trained on the full POAAGG training cohort (N = 271) for three feature configurations: Base, Base+PGS616, and Base+PC5+PGS616. A background summary was computed using shap.kmeans(X, k=20) to approximate the marginal feature distribution. SHAP values were estimated with nsamples=150 for computational efficiency. Mean absolute SHAP values (mean|SHAP|) were computed per feature across all training samples and used as the primary importance measure. SHAP beeswarm plots and bar charts are presented in Figure S3 and data is presented in Table S11.

### 9. Calibration Assessment

Model calibration was assessed using two complementary approaches. First, calibration curves (reliability diagrams) were generated using sklearn.calibration.calibration_curve with n_bins=10 and strategy=’uniform’ within 5-fold stratified cross-validation; predicted probabilities from all folds were pooled for visualization. Second, Brier scores were computed under the full 5×20 CV framework (100 fits per configuration) using sklearn.metrics.brier_score_loss.^62^ A null model Brier score (predicting the marginal outcome prevalence for all subjects) was computed as the reference (0.249 for the POAAGG training cohort). Calibration curves are presented for the MLP classifier (Figure S3C); Brier scores are reported for all four classifiers across three feature sets (Figure S3D).

### 10. Learning Curve Analysis

To assess whether model performance is sample-size limited, learning curves were generated using sklearn.model_selection.learning_curve with train_sizes = np.linspace(0.30, 1.00, 8) and 5-fold × 4-repeat stratified cross-validation (20 total fits per training size; random_state=42).^86^ Training AUC (on the training subset) and cross-validated AUC (on held-out folds) were computed using scoring=’roc_auc’. All four classifier pipelines were evaluated independently; classifier-averaged results are reported as the primary summary. Approximate absolute training set sizes were computed as the fraction multiplied by 80% × N (the typical per-fold training set size). Results are presented in Figure S4 and Table S13.

### 11. Sex-Stratified AUC Analysis

To evaluate sex-specific heterogeneity in model performance, sex-stratified cross-validation AUC was computed within the 5×20 CV framework (RepeatedStratifiedKFold, n_splits=5, n_repeats=20, random_state=42). Within each test fold, AUC was computed separately for male (Gender = 1) and female (Gender = 0) participants using sklearn.metrics.roc_auc_score. Folds with fewer than 5 samples of either sex or fewer than 2 outcome classes in a sex-stratified subset were excluded. The mean and standard deviation of the AUC across all valid folds (up to 100; folds not meeting the minimum sample criteria were excluded) are reported as descriptive summaries; confidence intervals and tests of the male–female difference come from the participant-level bootstrap (Table S19), which is also what Figure S5 displays. Results are reported for three feature configurations (Base, Base+PGS616, Base+PC5+PGS616) and summarized in Figure S5 and Table S14.

### 12. Prevention of Data Leakage

Every stage of the pipeline was checked for information flow from an evaluation cohort into model construction. A stage-by-stage summary, with the artifact demonstrating each safeguard, is given in Table S20.

### Feature selection

No data-driven feature selection was performed. The twelve feature sets were specified a priori from the study design, and every set was evaluated under identical procedures; no set was chosen on the basis of its performance.

### Polygenic score construction

The 271 participants in the machine learning training cohort were excluded from the POAAGG GWAS used to derive the POAAGG PGS and from the POAAGG component of the MEGA mega-analysis used to derive the MEGA PGS and the PGS616 weights, so no training participant contributed to any effect-size estimate used to score them. The curated variants were selected from external GWAS and were not screened against the training cohort. PMBB participants contributed to none of the score-derivation datasets.

### Hyperparameters

No hyperparameter search, grid search, or tuning of any kind was performed. All classifier settings were fixed a priori from published recommendations for imbalanced binary classification and were held constant across every feature set, cohort and analysis in the study. Consequently no evaluation data could influence model configuration.

### Preprocessing

Median imputation and z-score standardization were implemented as scikit-learn pipeline steps and were therefore refitted inside each cross-validation fold and each bootstrap replicate, using only the data in that fold’s or replicate’s training partition.

### Score standardization

The rank-based inverse normal transformation was applied separately in POAAGG (training and suspect cohorts pooled, N = 1,284) and in PMBB, so no distributional information passed between POAAGG and the external cohort. Within POAAGG the pooled transform uses no outcome labels, but the training and suspect cohorts are not standardized independently of each other.

### Ancestry principal components

The principal components were not derived independently of the modeling sample: they were computed once, on a set of POAAGG participants that includes both the modeling and suspect cohorts, and were not recomputed within folds or bootstrap replicates. The decomposition is unsupervised and used no case–control labels, so no outcome information entered the components, and the primary model contains no principal components; we state the dependence explicitly rather than claim independence.

### External and suspect cohorts

Models were trained once in the POAAGG training cohort and applied to the PMBB and suspect cohorts without refitting or recalibration. Neither cohort was used at any point to select models, features, or parameters. The suspect cohort contains no case–control labels and could not have been used for fitting. The one exception is the exploratory screening analysis, in which each model’s operating point was set at 90% or 95% specificity from the PMBB control distribution; these thresholds therefore use the external cohort’s labels and are not independently validated screening thresholds, although the same rule was applied to both models being compared.

## QUANTIFICATION AND STATISTICAL ANALYSIS

Model performance was quantified using standard classification metrics, including AUC, accuracy, F1-score, sensitivity (recall for the positive class), and specificity (recall for the negative class).^87^ Metrics were computed on held-out test sets across repeated stratified cross-validation iterations, and final reported values represent the mean and standard deviation across iterations. Brier scores were used to assess calibration, with the null model (marginal prevalence prediction) as reference.^62^ SHAP values were computed using KernelExplainer with a k-means background summary (k=20, nsamples=150).^61^ Learning curve analyses used train_sizes ranging from 30% to 100% of the per-fold training set under 5-fold × 4-repeat CV (20 fits per training-size step).^86^ Sex-stratified AUC was computed within each CV test fold and aggregated across 100 folds. Repeated cross-validation summaries report mean ± standard deviation and are descriptive. All 95% confidence intervals and p-values for the training cohort derive from a participant-level bootstrap (B = 2,000) in which participants are resampled with replacement, the pipeline is refitted on the in-bag sample and evaluated on out-of-bag participants; incremental comparisons are paired within replicate.

### Uncertainty estimation in the training cohort

Repeated cross-validation was used to characterize model stability, but its fold-level estimates are not statistically independent: the same 271 participants recur across all folds and repeats, so the standard error of the 100 fold-level AUCs understates the sampling variability of the cohort and cannot support interval estimation or hypothesis testing. We therefore report two distinct quantities and use only the second for inference.

### Descriptive

For each classifier and feature set we report the mean and standard deviation of the AUC across the 100 fits of 5-fold × 20-repeat stratified cross-validation. These summarize fit-to-fit stability and are reported without confidence intervals or p-values.

### Inferential

Uncertainty is estimated by a participant-level bootstrap with B = 2,000 replicates. In each replicate, participants are drawn with replacement from the training cohort, stratified on case–control status so that prevalence is preserved; the complete modeling pipeline (median imputation, z-score standardization, classifier) is refitted on the in-bag sample and evaluated on the out-of-bag participants, who contribute to no part of that replicate’s model fitting. The bootstrap is conditional on the constructed scores and components: the rank-based transformation of the polygenic scores and the ancestry principal components were computed once beforehand and are not re-estimated within replicates. Replicates leaving fewer than 10 out-of-bag members of either class are discarded. Reported 95% confidence intervals are percentile intervals of the resulting AUC distribution.

Because every feature set and every classifier is refitted on the same in-bag participants and evaluated on the same out-of-bag participants within a replicate, incremental comparisons between models are paired by construction, and the interval for an incremental AUC difference accounts for the correlation between the two models being compared. Two-sided p-values for incremental differences are computed as 2 × min(P(Δ ≤ 0), P(Δ ≥ 0)), bounded below by the reciprocal of the number of valid replicates. The same procedure is applied to the sex-stratified, inter-eye asymmetry and PGS-residualization analyses (Table S19), so that every incremental comparison reported for the training cohort rests on the same inferential basis.

External validation in PMBB uses participant-level bootstrap resampling (B = 2,000) of the validation cohort together with the DeLong test for two correlated ROC curves; models are trained once in the POAAGG cohort and applied without refitting, so no resampling of the training cohort is involved. Results are reported in Tables S6, S7, S15, S16 and S18.

Performance summaries for individual models and feature sets are presented in Figures 2–5 and Tables S5–S6. Given the nonparametric nature of the ML models and resampling-based evaluation framework, no assumptions of normality or homoscedasticity were required for ML model evaluation. Parametric assumptions underlying Pearson correlation and OLS regression in suspect enrichment analyses are considered approximately met given the large sample size (N=1,013).

## ADDITIONAL RESOURCES

This study is not a clinical trial; there is no clinical registry number. There are no external sites that have been generated to support discussion or use of the information/data/material created by the manuscript.

### Declaration of generative AI and AI-assisted technologies in the manuscript preparation process

During the preparation of this work, the authors used ChatGPT (OpenAI, GPT-5.2 and GPT-5.5) and Claude (Anthropic, Sonnet 4.6, Opus 4.8 and Opus 5) to assist with language refinement, clarity, and editorial revisions of the manuscript. The original analysis code was written independently by the authors; during the revision, these tools were also used to review the analysis code. All content generated with AI assistance was carefully reviewed, edited, and validated by the authors. The authors take full responsibility for the accuracy, integrity, and final content of the published article.

