## Supplemental Figures for "Multimodal Prediction of Primary Open-Angle Glaucoma Using Polygenic Risk Scores and Clinical Features in a High-Risk African Ancestry Cohort"

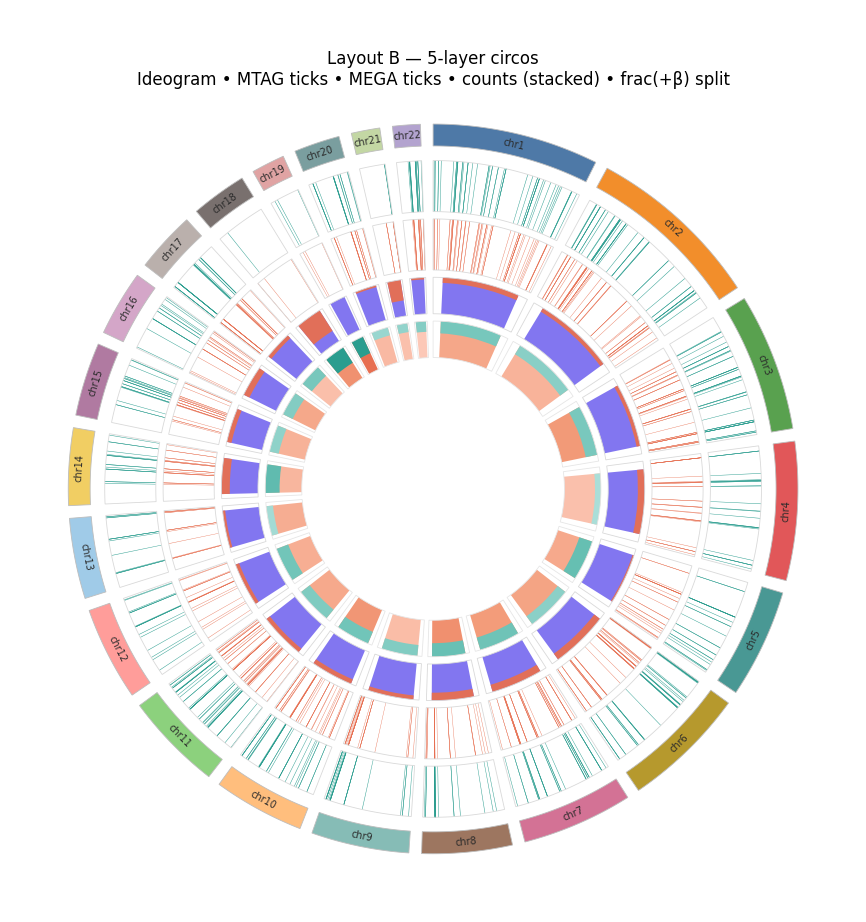

Figure S1. Chromosomal Distribution of Curated PGS Variants, Related to Figure 2

Circos plot showing the genomic distribution of the SNPs in the two curated locus-based polygenic scores across all 22 autosomes. The outermost ring displays the chromosomal ideogram with cytogenetic banding. The second ring (teal ticks) shows the positions of the 526 PGS526 variants, weighted by effect sizes from the POAAGG multi-trait analysis (MTAG); the third ring (orange ticks) shows the 616 PGS616 variants, weighted by MEGA African ancestry case–control effect sizes. PGS526 is a subset of PGS616, and both were drawn from genome-wide significant loci of multi-ancestry glaucoma GWAS (STAR Methods; Table S3). The fourth ring displays stacked variant density counts per chromosomal segment, and the innermost ring the fraction of positive-effect-size (β > 0) variants per segment, split by score. Variant density is highest on chromosomes 1, 2, 4, 6 and 9, and variants are spread across all 22 autosomes without focal clustering, so neither score is dominated by a small number of chromosomal regions.

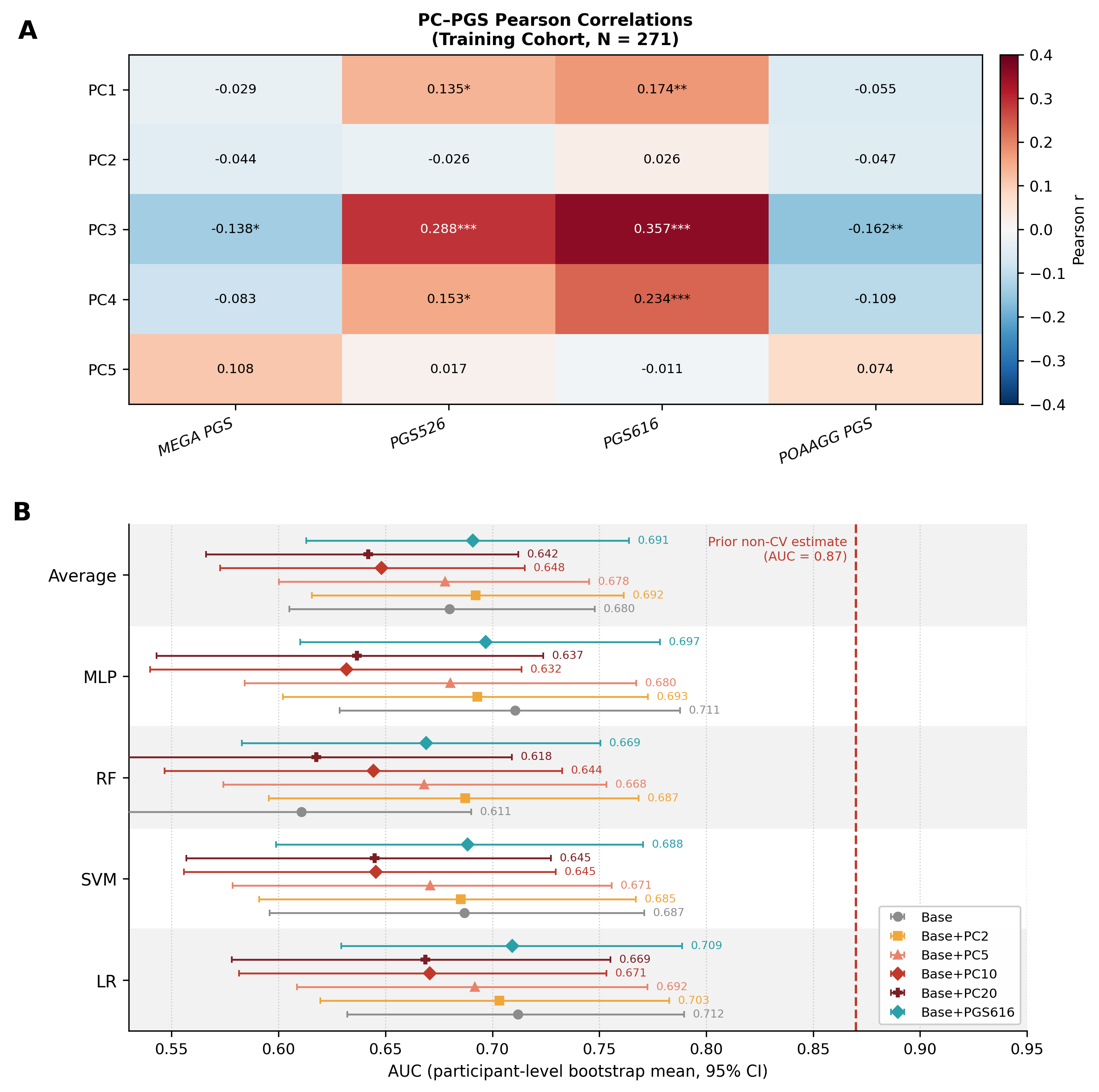

Figure S2. PC–PGS Correlations and Ancestry PC vs. PGS Performance, Related to Figure 3

(A) Heatmap of Pearson correlation coefficients between the first five ancestry principal components (PC1–PC5) and the four polygenic scores (MEGA PGS, PGS526, PGS616, POAAGG PGS) in the POAAGG training cohort (N = 271). Asterisks indicate statistical significance: *p < 0.05, **p < 0.01, ***p < 0.001. PC3 showed the strongest correlations with the curated loci-based scores (PGS526: r = 0.29; PGS616: r = 0.36; both p < 0.001), and modest negative correlations with MEGA PGS (r = −0.14, p < 0.05) and POAAGG PGS (r = −0.16, p < 0.01). PC1 showed weak positive correlations with PGS526 (r = 0.14, p < 0.05) and PGS616 (r = 0.17, p < 0.01); PC2, PC4 and PC5 showed weak or non-significant correlations. Because a correlation of 0.36 is not negligible, we do not interpret this as evidence that ancestry PCs and PGS capture distinct signals; a residualization analysis is reported in Tables S10 and S19. (B) AUC (participant-level bootstrap mean and 95% percentile confidence interval, B = 2,000) for six feature configurations — Base (age + sex), Base+PC2, Base+PC5, Base+PC10, Base+PC20 and Base+PGS616 — for each classifier (LR, RF, SVM, MLP) and their cross-classifier average. On the average, Base+PC2 was highest among the PC-augmented sets (0.692), with lower values as further components were added (Base+PC5 0.678, Base+PC10 0.648, Base+PC20 0.642), a pattern consistent with overfitting as dimensionality rises relative to N = 271; Base+PGS616 (0.691) was comparable to Base+PC2. The corresponding cross-validation means are in Table S5. None of these differences is statistically distinguishable, and no feature set differed significantly from the age and sex baseline (0.680; Table S8). The red dashed line marks AUC = 0.87, the value reported in a prior non-cross-validated analysis, retained to show how far that estimate overstated achievable performance.

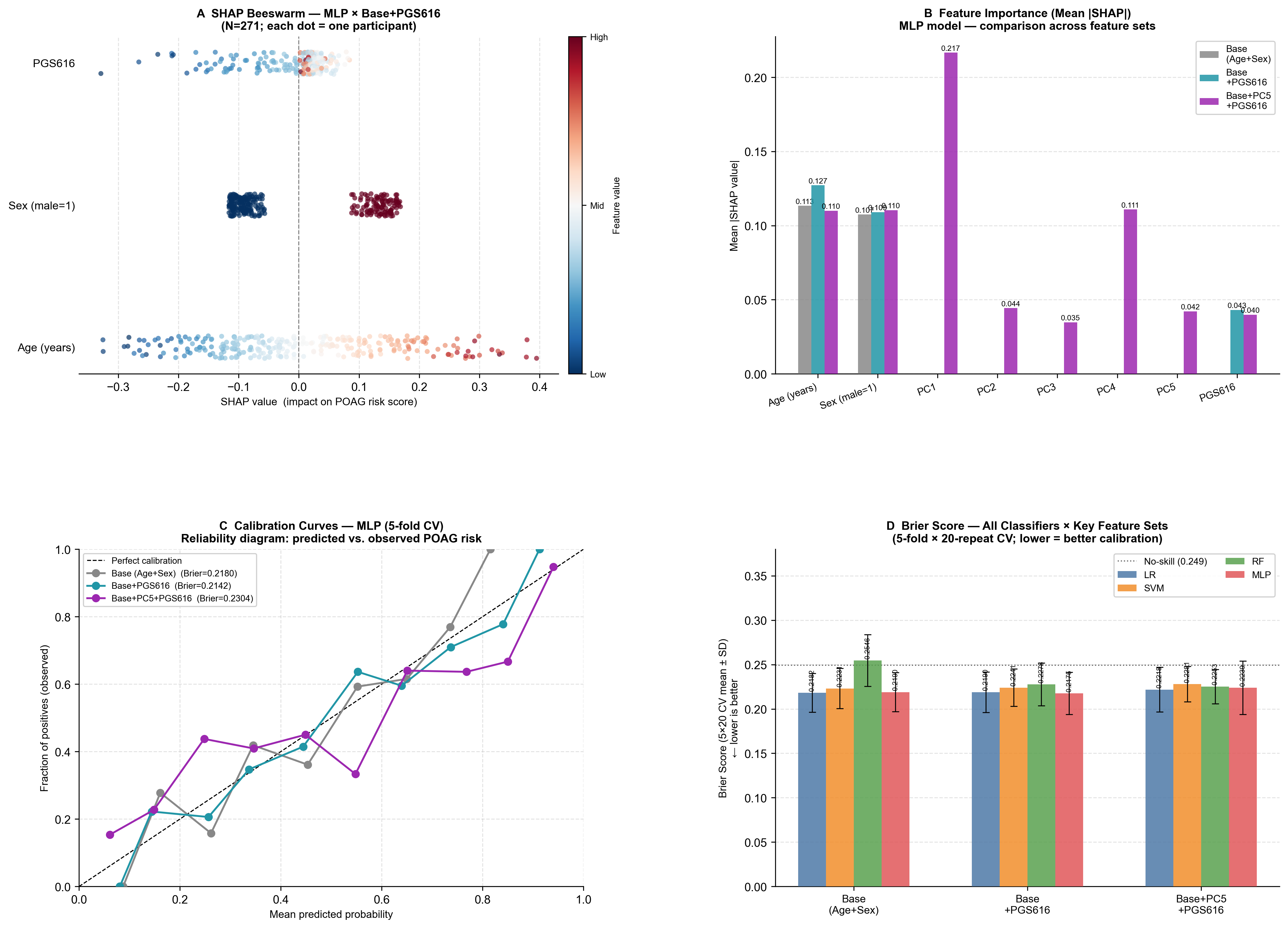

Figure S3. SHAP Feature Importance and Model Calibration, Related to Figure 3

(A) SHAP beeswarm plot for the MLP classifier trained on the Base+PGS616 feature set in the POAAGG training cohort (N = 271). Each dot represents one participant; the x-axis gives the SHAP value (impact on predicted POAG risk) and color the feature value (red = high, blue = low). Age is the most influential feature, with older age producing positive contributions; sex (male = 1) separates into two clusters, with male participants receiving higher predicted risk; PGS616 contributions are small, with higher values tending toward small positive contributions. (B) Mean |SHAP| for three MLP configurations: Base (age + sex), Base+PGS616 and Base+PC5+PGS616. For Base+PGS616, age (0.127) > sex (0.109) > PGS616 (0.043). When ancestry PCs are added (Base+PC5+PGS616), PC1 becomes the dominant feature (0.217), ahead of age (0.110), sex (0.110) and PGS616 (0.040). SHAP describes the fitted model and does not identify the biological source of a signal. (C) Calibration curves (reliability diagrams) for the three MLP configurations under 5-fold cross-validation; the dashed diagonal represents perfect calibration. Brier scores are shown in the legend: Base (0.2180), Base+PGS616 (0.2142), Base+PC5+PGS616 (0.2304). (D) Brier score (mean ± SD across the 100 fits of 5-fold × 20-repeat cross-validation; descriptive, not confidence intervals) for all four classifiers across the three feature sets. The dotted line at 0.249 is the expected Brier score of a constant prediction equal to the prevalence. Every combination except RF with Base (0.2546) lies below it. Base+PGS616 and Base are similar, and both are lower than Base+PC5+PGS616 for most classifiers; all differences are small relative to the fold-to-fold spread (Table S12).

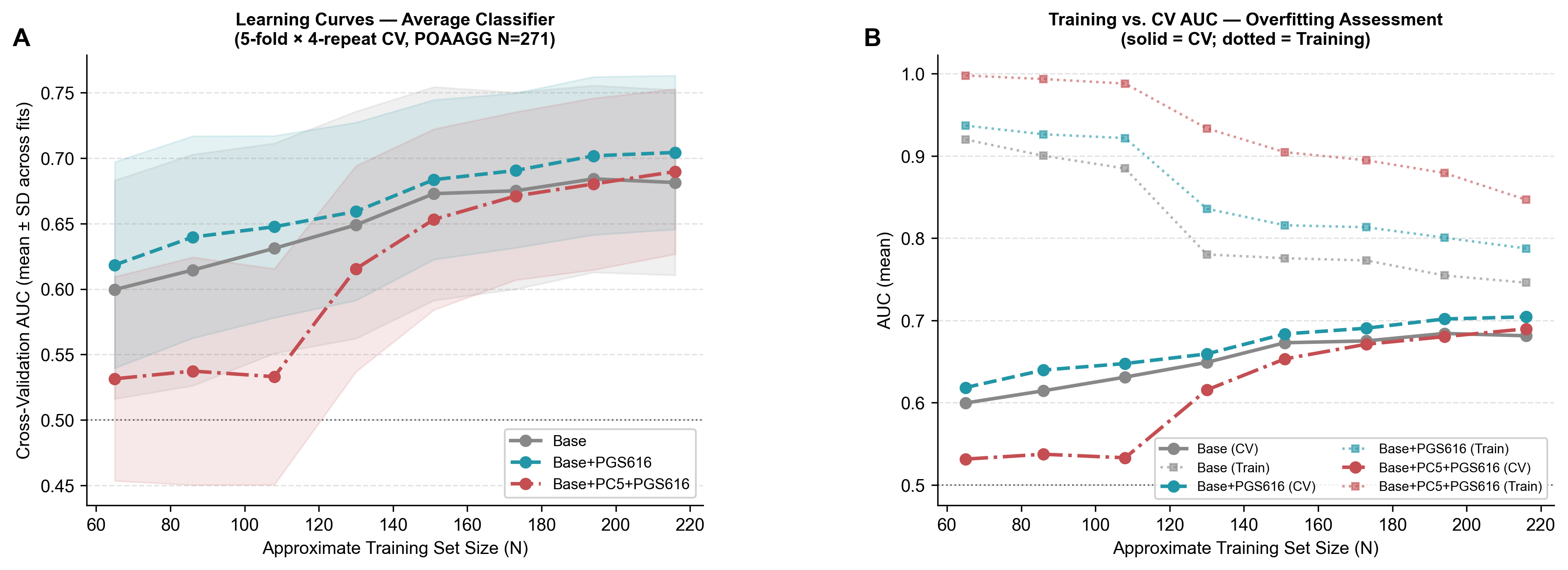

Figure S4. Learning Curves: Model Performance vs. Training Set Size, Related to Figure 3

(A) Learning curves showing cross-validation AUC (mean ± SD across the 20 fits of 5-fold × 4-repeat cross-validation; descriptive) as a function of approximate training set size for three feature configurations — Base (age + sex), Base+PGS616 and Base+PC5+PGS616 — averaged across the four classifiers. Training fractions ranged from 30% to 100% of the per-fold training set (approximate N = 65 to 216). Base+PGS616 (teal) lies above Base (gray) at every training size, although the curves are close relative to their spread. Base+PC5+PGS616 (red dashed) is much lower at small training sizes and approaches the other two only at full N, reflecting higher-dimensional overfitting at small sample sizes. Cross-validation AUC was still rising at the largest training size, so performance has not clearly plateaued. (B) Training AUC (dotted lines) versus cross-validation AUC (solid lines) for the three configurations, averaged across the four classifiers. At full training size the train-to-CV gap is 0.065 for Base, 0.083 for Base+PGS616 and 0.157 for Base+PC5+PGS616, whose training AUC is close to 1.0 at small N; adding ancestry PCs therefore increases overfitting, particularly at smaller sample sizes (Table S13).

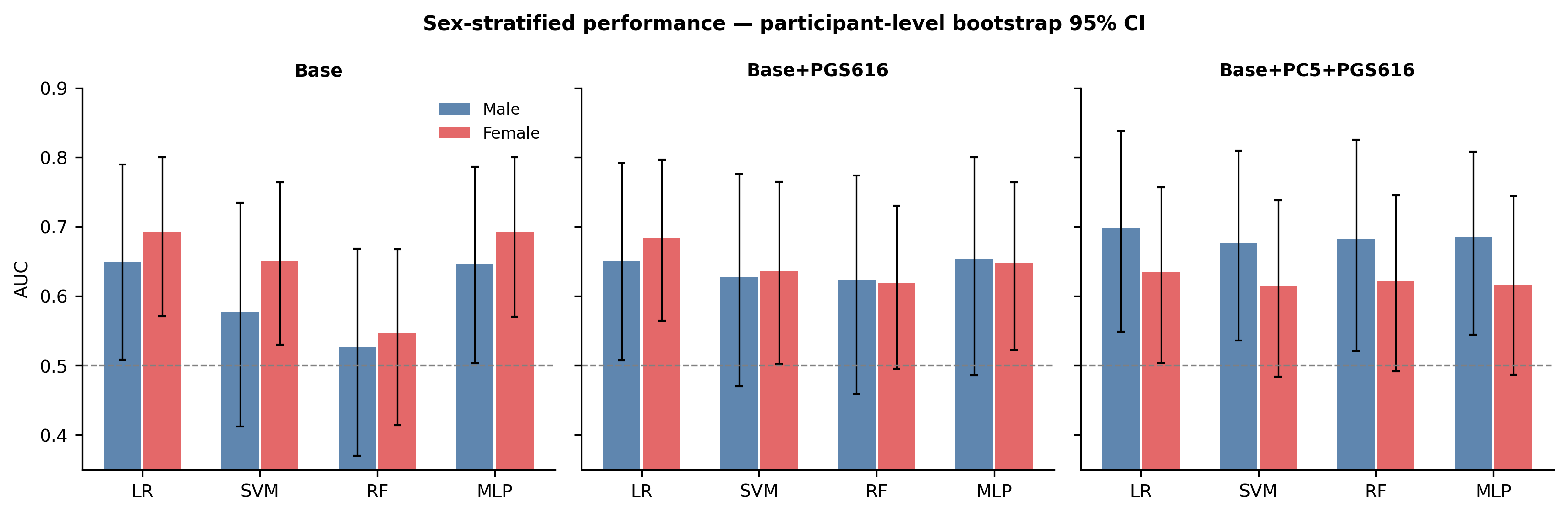

Figure S5. Sex-Stratified AUC, Related to Figure 3

AUC by sex for each classifier (LR, SVM, RF, MLP) and three feature configurations — Base (age + sex), Base+PGS616 and Base+PC5+PGS616 — in the POAAGG training cohort (male: N = 111, 68 cases; female: N = 160, 60 cases). Bars are participant-level bootstrap estimates and error bars 95% percentile confidence intervals (B = 2,000); the dashed line marks chance. For the primary MLP model, females had higher AUC than males with Base (0.691 versus 0.646), the two were similar with Base+PGS616 (0.647 versus 0.653), and males were nominally higher with Base+PC5+PGS616 (0.684 versus 0.616). No male–female difference was statistically distinguishable for any classifier or feature set (all p ≥ 0.47; Table S19), so these patterns are suggestive only. Descriptive cross-validation means by sex are given in Table S14.

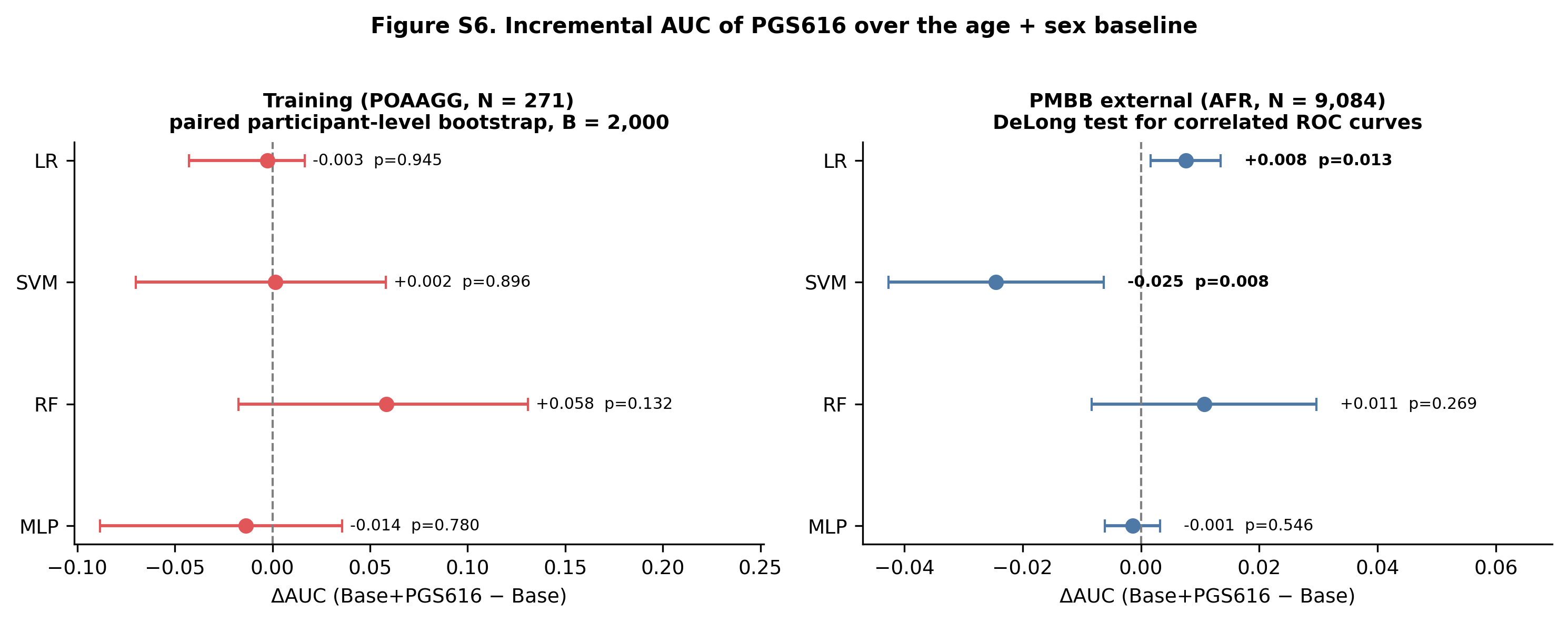

**Figure S6. Incremental AUC (ΔAUC) of PGS616 over the Age + Sex Baseline, Related to Figure 3**

Forest plot of the incremental AUC (ΔAUC = (Base+PGS616) − Base) for each classifier (LR, SVM, RF, MLP), computed within the same classifier. Left: training cohort (POAAGG, N = 271), paired participant-level bootstrap (B = 2,000); within each replicate both models are fitted on the same in-bag participants and evaluated on the same out-of-bag participants, and the bars are 95% percentile intervals. Right: PMBB external validation (AFR, N = 9,084), DeLong test for two correlated ROC curves with its analytic 95% interval; the same estimates, together with paired participant-level bootstrap intervals, are reported in Table S7. The dashed line marks ΔAUC = 0 (no improvement over the age + sex baseline). Points to the right of zero indicate that adding PGS616 improves discrimination; intervals overlapping zero indicate no statistically significant incremental gain. Across classifiers the incremental value of PGS616 is small and its sign varies; for the primary MLP model it is small in both cohorts (external ΔAUC = −0.001, DeLong p = 0.55).

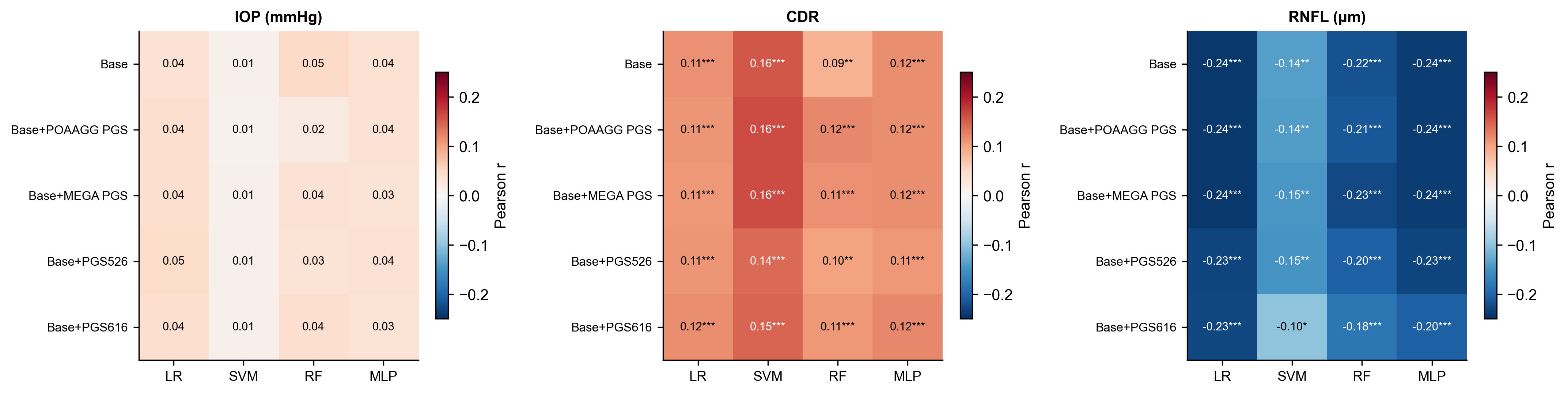

Figure S7A. Predicted Risk Correlations with Clinical Outcomes Across All Classifiers, Related to Figure 4

Heatmap of Pearson correlation coefficients between model-predicted POAG risk and three clinical measures — intraocular pressure (IOP), cup-to-disc ratio (CDR) and retinal nerve fiber layer (RNFL) thickness — in the POAAGG suspect cohort (N = 1,013), for all four classifiers (LR, SVM, RF, MLP) and five feature sets (Base, Base+POAAGG PGS, Base+MEGA PGS, Base+PGS526, Base+PGS616). Color represents the correlation coefficient (red = positive, blue = negative); asterisks denote statistical significance (*p < 0.05, **p < 0.01, ***p < 0.001). CDR was positively and RNFL thickness negatively associated with predicted risk in every classifier and feature set, while IOP associations were weak and non-significant throughout. The associations are as strong for the Base model, which contains no polygenic score, as for the PGS-augmented models, and after adjustment for age and sex the two curated scores tested (PGS616 and PGS526) have small, non-significant associations with each phenotype (Table S17): the enrichment is attributable to the age and sex components of the models.

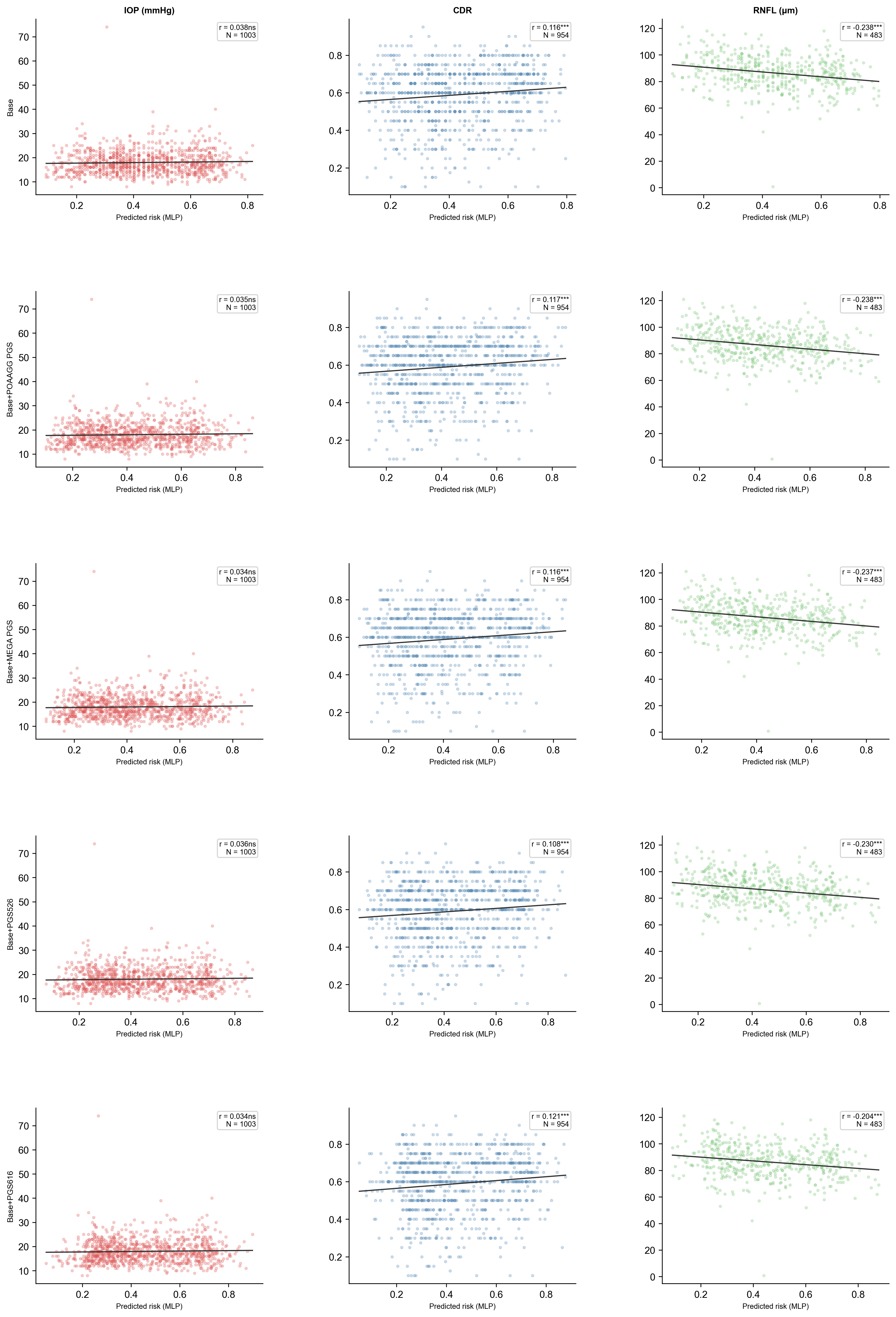

Figure S7B. MLP Predicted Risk vs. Clinical Outcomes in the Suspect Cohort, Related to Figure 4

Scatter plots of MLP-predicted POAG risk (x-axis) against intraocular pressure (IOP, mmHg), cup-to-disc ratio (CDR) and retinal nerve fiber layer (RNFL) thickness (μm) in the POAAGG suspect cohort, for five feature sets (Base, Base+POAAGG PGS, Base+MEGA PGS, Base+PGS526, Base+PGS616). Regression lines with 95% confidence bands are shown; correlation coefficients and sample sizes are displayed in each panel. IOP showed no significant association with predicted risk in any feature set (r = 0.034–0.038, all p > 0.2; N = 1,003). CDR showed significant positive correlations in every feature set (r = 0.108–0.121, all p < 0.001; N = 954), and RNFL thickness significant negative correlations (r = −0.204 to −0.238, all p < 0.001; N = 483). The RNFL association is r = −0.238 for the age-and-sex model and r = −0.204 with PGS616 added; after adjustment for age and sex the two curated scores tested (PGS616 and PGS526) have small, non-significant associations with each phenotype (Table S17), so these correlations reflect the demographic components of the models.
